# Anti-CELA1 KF4 Antibody Prevents Emphysema by Inhibiting Stretch-Mediated Remodeling

**DOI:** 10.1101/2020.04.27.20075531

**Authors:** Mohit Ojha, Noah Smith, Rashika Joshi, Emily Goodman, Qiang Fan, Richard Schuman, Aleksey Porollo, Rachel L. Zemans, Matthew R. Batie, Michael T. Borchers, Samuel Ammerman, Brian M. Varisco

## Abstract

Emphysema is a major contributor to the morbidity and mortality of chronic obstructive pulmonary disease (COPD), and there are no disease modifying therapies. Three leading pathophysiologic models are the altered protease/antiprotease balance model, the biomechanical model in which loss of one alveolar wall increases strain on adjacent wall predisposing them to failure, and the accelerated aging model. *Chymotrypsin-like elastase 1* (*CELA1*) is a novel serine protease with a physiological role in reducing postnatal lung elastance that mechanistically links these three models. *CELA1* is expressed by alveolar type 2 (AT2) cells and is found adjacent to lung elastin fibers. The KF4 antibody neutralizes CELA1 by binding to its catalytic triad. CELA1 binding to lung elastin fibers increases 4-fold with strain, and application of biaxial strain induces lung elastase activity which is blocked with the KF4 anti-CELA1 antibody. *Cela1^−/−^* mice were protected from airspace simplification in hyperoxia, elastase and cigarette-smoke induced emphysema, and age-related airspace simplification. *CELA1* mRNA was correlated with human lung elastolytic activity, and anti-CELA1 KF4 antibody protected mice from hyperoxia-induced alveolar simplification and elastase emphysema. CELA1-mediated lung matrix remodeling in response to strain is an important contributor to postnatal airspace simplification. Matrix stabilization by KF4 represents a potential therapeutic approach to preventing emphysema progression.

**One Sentence Summary:** Neutralization of chymotrypsin-like elastase 1 prevents strain-induced emphysema.

## Introduction

The lung is a complex and delicate structure with a surface area of about 150 m^2^ through which gas exchange occurs. This surface area is accommodated by the thoracic cavity via a highly stereotyped pattern of branching airways terminating in thin alveolar sacs which are septated and perfused by an extensive capillary network. Emphysema is the term used for loss of these alveolar structures with resultant decrease in gas exchange capacity. Loss of alveoli is termed alveolar simplification or emphysema and is an element of many respiratory diseases such as bronchopulmonary dysplasia, connective tissue disorders, alpha-1 antitrypsin (AAT) deficiency, and chronic obstructive pulmonary disease (COPD). COPD is the most common cause of emphysema, accounting for about $50 billion in US health care expenditures annually. Before the COVID-19 pandemic, COPD was the 3^rd^ leading cause of death worldwide (*1*). While a host of immune, epithelial, stromal, and vascular cell defects have been described, an important defect in COPD is loss of lung matrix (*2*). This can be observed quite strikingly in comparing lung computed tomography images of healthy and COPD subjects in which the later have loss of large areas of matrix and associated cells (*3*).

Among the host of extracellular matrix components in the lung, none is perhaps more important that elastin. Elastin fibers are composed of a host of elastin-associated proteins and a core of tropoelastin monomers which are hydrophobic and arranged in a repetitive overlapping pattern (*4*). When tension is applied to elastin fibers, potential energy is stored as these hydrophobic domains are separated giving lung its elastic properties which are key to passive exhalation. While many proteases have been shown to degrade lung elastin fibers, we have previously described a unique role for *Chymotrypsin-like Elastase 1* (*CELA1*) in alveolar remodeling (*5*). CELA1 is a serine protease that has traditionally been considered a digestive enzyme but has a developmentally regulated expression pattern in AT2 cells (*6*). In the lung, *CELA1* expression is increased with strain (*7*). Like other pancreatic elastases, CELA1 binds to hydrophobic domains of tropoelastin when exposed by stretch (*7*). The normal role for CELA1 is to reduce postnatal elastance, and *Cela1^−/−^* mice are phenotypically normal except for having higher lung elastance and smaller alveoli than wild type mice (*5*). CELA1 is neutralized by α1-antitrypsin (AAT) and is required for development of emphysema in a mouse model of AAT-deficient emphysema (*6*).

Multiple models of emphysema pathogenesis exist. The altered protease/antiprotease balance model developed after the discovery of α1-antitrypsin by Laurell and Erikson in 1964 (*8*) holds that unopposed protease activity leads to tissue destruction and emphysema. The accelerated aging model is based on similarities in airspace architecture between COPD and aged human lung and similar change in cellular state, oxidative stress and matrix architecture (*9*). The biomechanical model holds that strain is distributed over alveolar walls and associated matrix structures and that alveolar wall disruption increases strain on adjacent walls and predisposes them to failure. This manuscript details how *CELA1* mechanistically unites these three models and promotes a feed-forward process of alveolar destruction.

Given the stretch-dependent nature of CELA1 in lung elastin remodeling, we hypothesized that *CELA1* mediates degradation of lung elastin fibers in regions of the lung that experience localized areas of increased strain. Such a model has been predicted and validated in simulated and animal models (*10–13*), and is termed the biomechanical model of emphysema. In this model, loss of alveolar structure imparts additional stress on remaining structures leading to progressive airspace destruction. Here, we demonstrate that *CELA1* plays an important role in emphysema progression in multiple models and at multiple stages of the life cycle, that CELA1 binds to lung elastin fibers after stretch and mediates stretch-induced lung elastase activity in healthy lung tissue. Inhibiting CELA1 with the KF4 antibody represents a potential approach to preventing emphysema progression in multiple disorders of alveolar simplification.

## Results and Discussion

### In the Adult Lung, CELA1 is Expressed in Surfactant Protein C-High Alveolar Type 2 Cells

We previously identified CELA1 protein in lung epithelial, immune, and mesenchymal lung cells (*5*) but noted that *Cela1* mRNA was restricted to distal lung epithelial cells in the mouse lung and highest at E18.5 shortly before birth (*7*). We queried mouse and human lung cell datasets to better define *CELA1*-expressing cells (*14, 15*). The Choi, *et al* dataset (GSE145031) assessed lung cell expression in the murine porcine pancreatic elastase (PPE) model. As reported, alveolar type 2 (AT2) cells clustered in surfactant protein C (Sftpc)-high and Sftpc-low clusters (Figure 1A-C). *Cela1* expression was almost entirely restricted to Sftpc-high cells. Sftpc-high cells represented ~60% of total AT2 cells while in PPE-treated lung they represented ~40% (Figure 1D-E). AT2 cell expressing *Cela1* had higher mRNA levels of genes related to oxidative metabolism (Figure 1F). In human lung single cell analysis of control, idiopathic pulmonary fibrosis (IPF) and COPD subjects (GSE136831), *CELA1* mRNA was also restricted to SFTPC-high cells; although, unlike the mouse experiment, a subset of AT2 cells with pro-inflammatory genes was observed (Figure 1G-I). Most AT2 cells in IPF and COPD lung were SFTPC-low, and almost 75% of AT2 cells in control lung were SFTPC-high (Figure 1J). Five and two percent of AT2 cells were *CELA1* mRNA-positive in control and IPF or COPD lung respectively (Figure 1K). *CELA1-*expressing human AT2 cells also had enrichment for oxidative phosphorylation-related mRNAs (Figure 1L) demonstrating similarity between mouse and human CELA1-expressing AT2 cells. In mouse lung (Figure 1M) and human lung (Figure 1N), *CELA1* mRNA and protein were present in AT2 cells. *CELA1* is expressed in a similar subset of AT2 cells in mouse and human lung.

**Figure 1:**
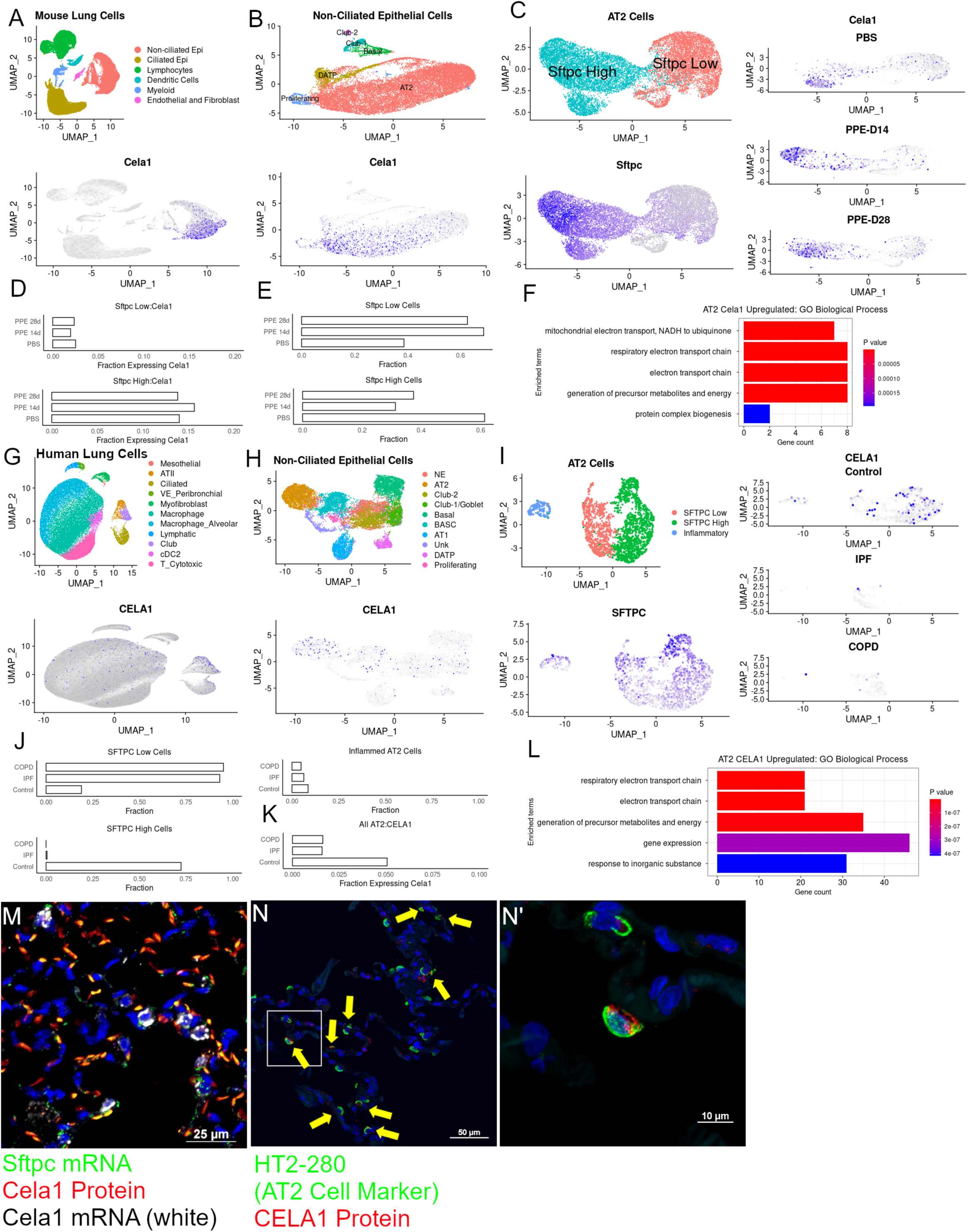
Single Cell Analysis of CELA1-expressing Cells in Lung. (A) A Uniform Manifold Approximation and Projection (UMAP) of mouse lung cells from a published experiment in which mice were treated with PBS or PPE with lungs collected at 14 and 28 days. Major cell types are shown (top) and a *Cela1*-expressing cells are highlighted in purple on the bottom. These were restricted to non-ciliated epithelial cells. (B) Sub-setting and re-clustering of non-ciliated epithelial cells identified the expected cell types (top) with *Cela1* expression in non-proliferating, non-differentiating AT2 cells (bottom). (C) Sub-setting and re-clustering of AT2 cells identified the previously described Sftpc-high and -low clusters. Cela1-expressing cells were largely restricted to Sftpc High AT2 cells in all three treatment groups (right). (D) A barplot of the percent Sftpc Low and Sftpc High cells with *Cela1* mRNA showing ~2% and ~15% expression regardless of treatment group. (E) In PPE-treated lungs, ~60% of cells were Sftpc Low compared to 40% in PBS-treated lungs indicating a shift in cell population distribution after PPE. (F) In comparing the gene expression of *Cela1*-expressing vs Non-*Cela1*-expressing AT2 cells, mRNAs of oxidative phosphorylation genes were enriched. (G) UMAP of human lung cells from a dataset of non-lung organ donor, end stage COPD, and end stage IPF lungs showing the expected cell types (top) with CELA1 expression in non-ciliated epithelial cells. (H) Sub-setting and re-clustering of non-ciliated epithelial cells again identified expected cell types (top) with CELA1 expressing cells mostly in AT2 cells (bottom). (I) Sub-setting and re-clustering of human AT2 cells again identified SFTPC High and SFTPC Low clusters but also a cluster enriched for inflammatory genes. CELA1-mRNA-containing cells were mostly in the SFTPC-high cluster which was largely absent from COPD and IPF lung. (J) Bar plot showing that ~90% of AT2 cells in end stage IPF and COPD lung were SFTPC-low compared to ~15% in organ donor lung (Control). Inflammatory AT2 cells comprised about 5% in each group. (L) Comparing differentially expressed genes in *CELA1*-expressing vs. non-*CELA1*-expressing AT2 cells, showed enrichment of oxidative phosphorylation genes. (M) In mouse lung, *Cela1* mRNA (white) and protein (red) colocalized with *Sftpc* mRNA positive cells, although some *Sftpc*-expressing cells did not have Cela1 mRNA or protein. Scale bar = 25 µm. (N) In human lung, a subset of AT2 cells (HT2-280 positive, green) contained CELA1 protein (red). Scale bar = 50 µm. (N’) Magnification of panel N. Scale bar = 10 µm. Epi=Epithelial Cells, AT2=Alveolar Type 2 Cells, DATP=Damage-Associated Transient Progenitor Cells, Sftpc=Surfactant Protein C, Cela1=Chymotrypsin-like Elastase 1, PPE=Porcine Pancreatic Elastase, COPD=Chronic Obstructive Pulmonary Disease, IPF=Idiopathic Pulmonary Fibrosis

### CELA1 Expression in the Injured Mouse Lung

As reported previously, *Cela1* is expressed at a high level in distal lung epithelial cells at E18.5 (Supplemental Figure 1A). *Cela1* expression is increased in post-pneumonectomy lung in response to strain (*7*). We quantified lung mRNA levels in three different mouse models of lung injury. Following PPE administration, lung *Cela1* mRNA levels were variable at days 1-21 but significantly higher at 42 days compared to PBS (Supplemental Figure 1B). In the neonatal hyperoxia model, lung *Cela1* mRNA levels were almost twice that of room air control (Supplemental Figure 1C). In the cigarette smoke exposure model, *Cela1* mRNA levels increased with increasing duration of exposure (Supplemental Figure 1D). As each of these three models are associated with progressive airspace simplification, there is an association of increasing *Cela1* expression with increasing airspace remodeling.

### CELA1-binding and Stretch-Inducible Elastase Activity in Human Lung

In human lung, CELA1 was present in the extracellular matrix in association with elastin fibers (Figure 2A). CELA1 binds and cleaves non-crosslinked tropoelastin at hydrophobic domains, and we hypothesized that like other pancreatic elastases (*16*), CELA1 binds and cleaves tropoelastin when hydrophobic domains are exposed by strain. In mouse lung, strain increased CELA1 binding to lung elastin increased ~4-fold (*7*). In human lung, with increasing levels of biaxial strain the binding of fluorophore-conjugated CELA1 but not albumin increased (Figure 2B-E, Supplemental Figure 2). Biaxial stretch induces lung elastase activity in wildtype but not *Cela1^−/−^*mouse lung sections (*6*). In human lung sections, biaxial strain also increased lung elastase activity (Figure 2F-H) suggesting that similar strain-related remodeling programs are operative in human and mouse lung.

**Figure 2:**
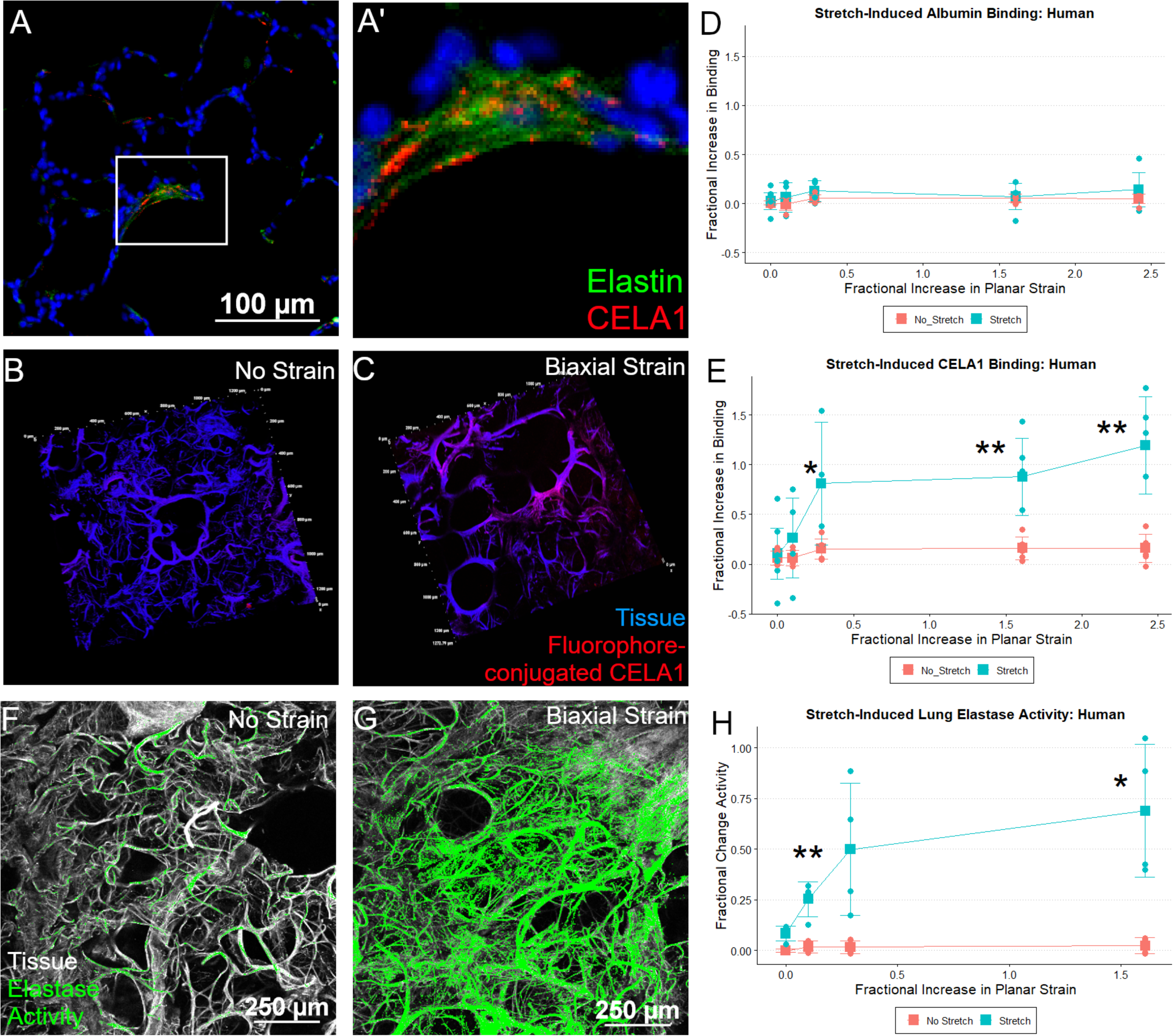
CELA1 and Stretch-Inducible Elastase Activity in Human Lung. (A) Immunofluorescence imaging for human tropoelastin (green) and CELA1 (red) demonstrated that in addition to its presence in AT2 cells, CELA1 protein can be found near elastin in the airspace walls. Scale bar = 100 µm. (A’) Higher magnification image of B. (B) Using fluorophore-conjugated CELA1, human lung tissue sections demonstrated little binding of CELA1 (red) to lung tissue (blue) in the absence of strain. Tissue section is 100 µm thick and each tick mark represents 200 µm. (D) When subjected to biaxial strain, CELA1 binding to lung tissue increased. (D) Fluorophore-conjugated albumin binding to lung tissue did not increase with strain. (E) Quantification of CELA1 binding in four independent lung specimens shows that binding increases with increasing levels of strain to about 5-fold. Comparisons by Welch’s t-test. (F) Using a fluorophore-conjugated and quenched soluble elastin substrate for *in situ* zymography, human lung tissue (white) does not have much elastase activity (green) at baseline. Scale bar = 250 µm. (G) When subjected to biaxial strain, there is an increase in lung elastase activity. (H) Lung elastase activity increases with strain by about 8-fold.

### Human Lung CELA1 mRNA Levels Correlate with Elastolytic Activity

We quantified *CELA1* mRNA and protease activity in lung homogenates from 23 non-lung organ donors and 8 COPD subjects. *CELA1* mRNA levels varied logarithmically but were higher in COPD with emphysema specimens compared to control (Figure 3A). Similar patterns were observed with *Matrix Metalloproteinase-8* (*MMP8*), *MMP9*, *MMP12*, *MMP14*, *Proteinase-3*, *Cathepsin G*, *Neutrophil Elastase*, *Tissue Inhibitor of Matrix Proteinase-1* (*TIMP1*), *TIMP2*, *TIMP1*, and *α1-Antitrypsin* (*SERPINA1*), but not *MMP2* (Supplemental Figure 3). Lung elastase, protease, and gelatinase activities were lower in COPD specimens compared to organ donor specimens (Figure 3B, Supplemental Figure 3). To investigate this apparent contradiction, we compared gene mRNA levels with protease, elastase, and gelatinase activities in each specimen. Whereas the mRNA levels of most proteases and antiproteases positively correlated with each other, *CELA1* mRNA levels were not positively correlated with those of other proteases, and only *CELA1* mRNA levels positively and significantly correlated with lung elastase and protease activity levels (Figure 3C).

**Figure 3:**
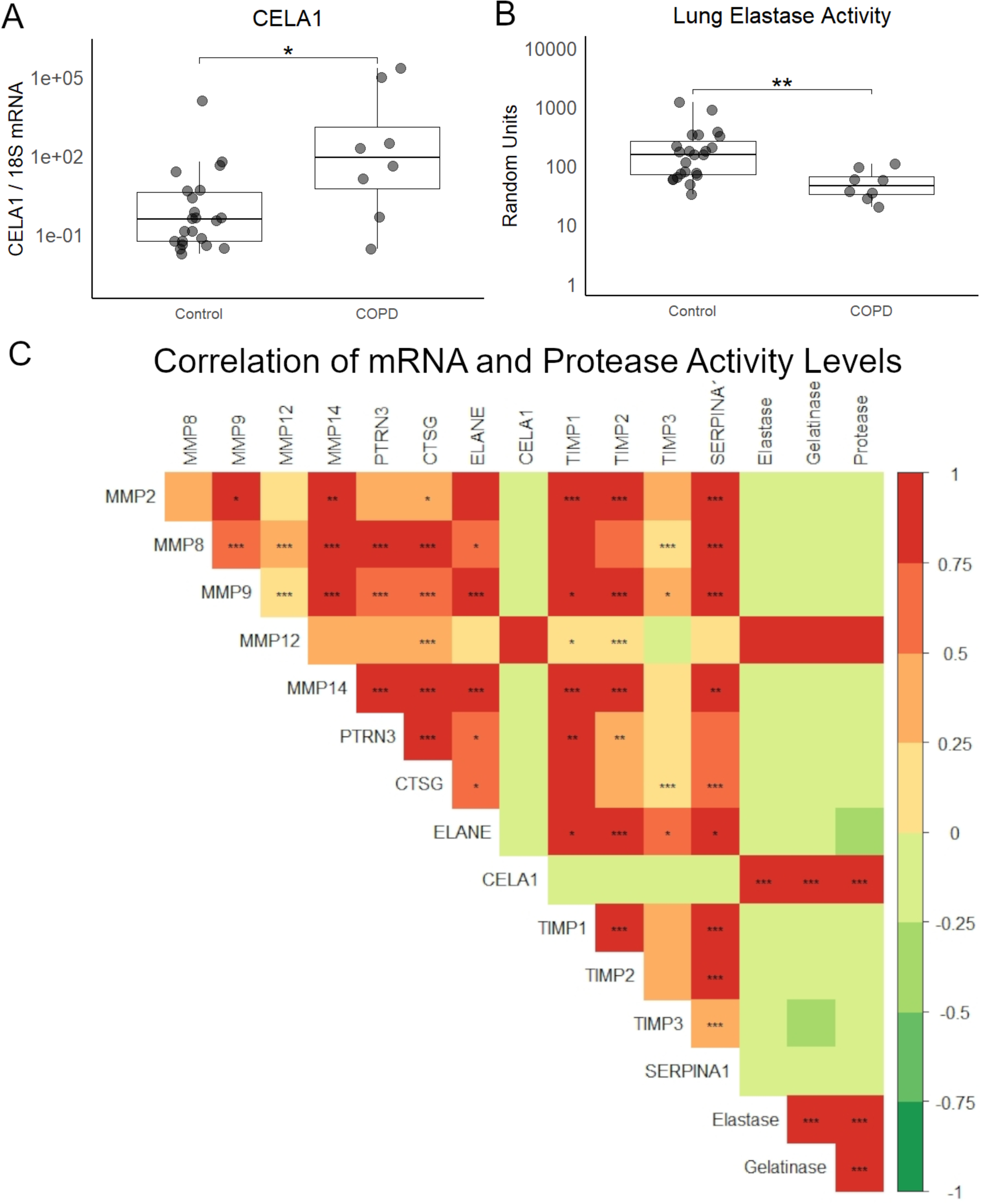
CELA1, Other Proteases, and Human Lung Elastase Activity. (A) In 14 non-smoker, 9 smoker, and 8 COPD lung homogenates, CELA1 mRNA levels were variable but overall higher in COPD lung than smoker control but not different than nonsmoker (NS) control. Kruskal-Wallis p<0.01, Dunn post hoc comparison shown. (B) In comparing the elastase activity of these homogenates, COPD homogenates had lower levels of elastase activity than NS control. Kruskal-Wallis p<0.01, Dunn *post hoc* comparison shown. (C) mRNA levels of *matrix metalloproteinase 2 (MMP2), MMP8, MMP9, MMP12, MMP14, Proteinase-3, Cathepsin G, Neutrophil Elastase, CELA1, Tissue Inhibitor of Metalloproteinase-1 (TIMP1), TIMP2, TIMP3, and α1-antitrypsin (SERPINA1)* as well as tissue protease, gelatinase, and elastase activities were compared by Spearman rank correlation with Bonferroni correction for multiple comparisons. *CELA1* and *MMP12* mRNA levels correlated with each other and with tissue protease, elastase, and gelatinase levels. The other proteases and anti-proteases were generally correlated with each other. *p<0.05, **p<0.01, ***p<0.001

### CELA1 Genomics

The gene *CELA1* was not previously identified in COPD genome wide association studies (*17, 18*). *CELA1* is highly conserved in placental mammals (*6*), and *CELA1* mutations in humans are rare with the most common predicted loss of function mutation having an allele frequency of 0.03% (Table 1) (*19–21*), and no predicted gain-of-function mutations are known (*22, 23*). As homozygous loss of function would be protective and occur in 1:11.7 million individuals, COPD GWAS studies would have been underpowered to identify an association of the *CELA1* locus with disease.

**Table 1:** Frequency of CELA1 Mutations

| <u>Position</u> | <u>RSID</u> | <u>Reference</u> | <u>Alternate</u> | <u>Protein Consequence</u> | <u>Annotation</u> | <u>Functional Prediction</u> | <u>Allele Frequency</u> |
| --- | --- | --- | --- | --- | --- | --- | --- |
| 51737562 | rs17860300 | T | C | p.Met59Val | missense | Benign | 0.2376 |
| 51723598 | rs60311818 | A | AG | p.Leu210ProfsTer24 | frameshift | Benign | 0.2295 |
| 51737607 | rs17860299 | G | A | p.Arg44Trp | missense | Benign | 0.1982 |
| 51739648 | rs17860287 | C | G | p.Gln10His | missense | Benign | 0.09864 |
| 51723499 | rs17860364 | T | C | p.Gln243Arg | missense | Benign | 0.09134 |
| 51739640 | . | G | A | p.Pro13Leu | missense | Benign | 0.00909 |
| 51739625 | rs74336876 | C | T | p.Arg18His | missense | Benign | 0.007377 |
| 51735144 | rs17860313 | C | T | p.Arg116His | missense | Benign | 0.001723 |
| 51735071 | . | AG | A | p.Ala140ValfsTer47 | frameshift | Benign | 0.0005204 |
| 51733775 | . | C | T | p.Ala160Thr | missense | Loss-of-Function | 0.0002924 |

\* Compilation of data from dbGap, ClinVar, and Broad Institute. Predictions by SIFT and PolyPhen-2.

### *Cela1* Deficient Mice Have Preserved Alveolar Architecture in Multiple Models of Airspace Simplification

To test whether *CELA1* has a general role in airspace simplification, we tested *Cela1*-deficient mice in multiple models. *Cela1^−/−^* mice were protected from airspace simplification in the hyperoxia model of impaired alveolar development (Figure 4A, Supplemental Figure 4), from emphysema progression after intratracheal administration of PPE (Figure 4B, Supplemental Figure 5), from emphysema following 8 months of cigarette smoke exposure (Figure 4C, Supplemental Figure 6), and from age-related alveolar simplification (Figure 4D). The lungs of aged *Cela1^−/−^* mice had preserved elastin bundles at alveolar tips as assessed by Hart staining and brightfield microscopy, and greater total lung elastin by Western blot (Figure 4E). Although we previously reported that *Cela1* mRNA and protein were increased in the mouse partial pneumonectomy model of lung regeneration (*7*), we found no difference in compensatory lung regrowth at 7 or 28 days (Supplemental Figure 7). Sex was not a substantial factor in these analyses (Supplemental Figures 4–7). Taken together, these data show a central role for *CELA1* in alveolar destruction and no role in regeneration.

**Figure 4:**
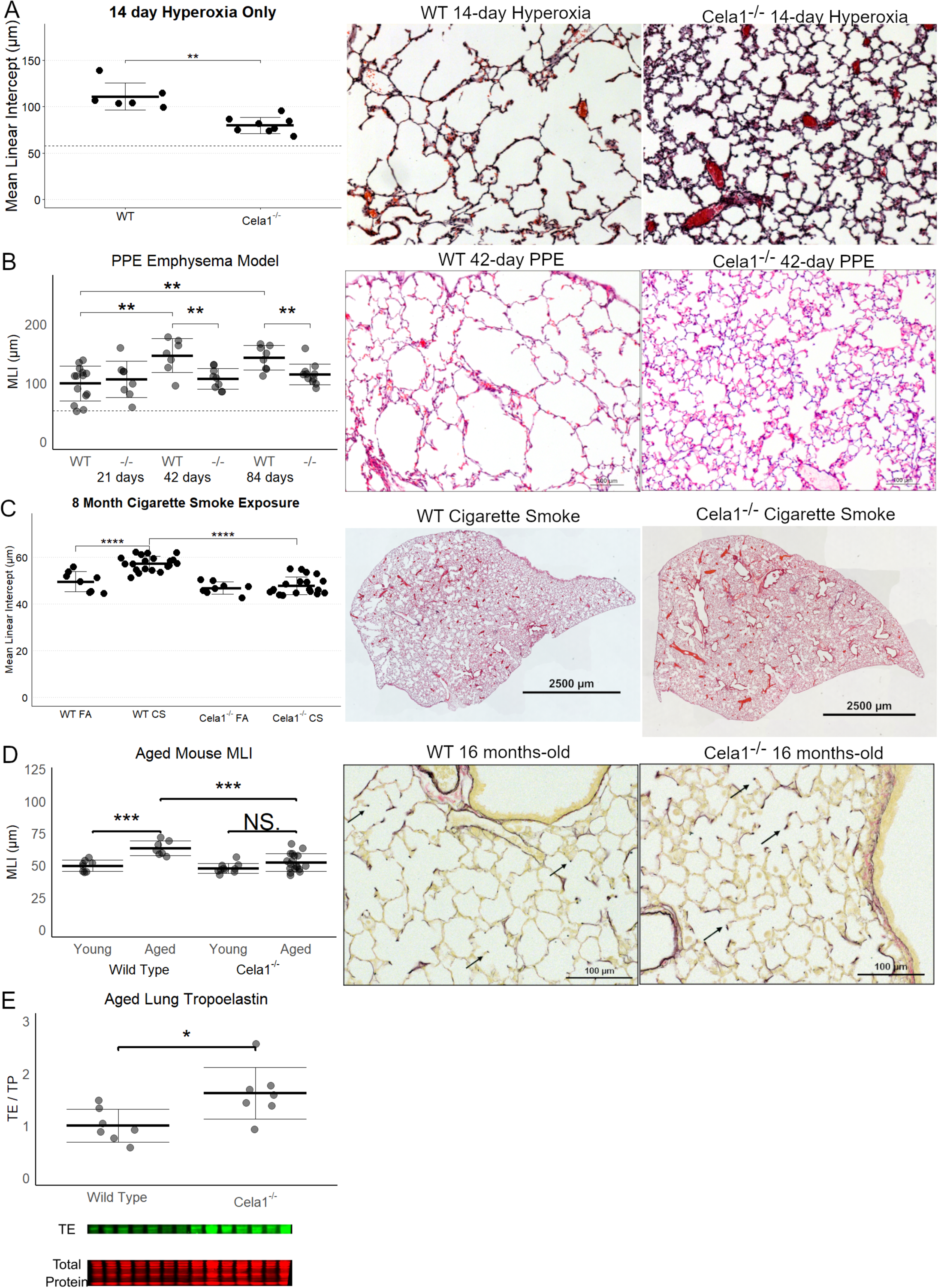
Protection of Cela1-/- Mice in Multiple Models of Alveolar Simplification. (A) As assessed by mean linear intercept (MLI), *Cela1^−/−^* mouse pups (n=8) had about half of the airspace simplification of wildtype (WT, n=6) mice after exposure to 80% oxygen for the first 14 days of life. Dashed line represents average MLI of PND14 WT mice exposed to room air. Comparison by Welch’s t-test. Representative 10X photomicrographs are shown. (B) At 21-days after airway administration of porcine pancreatic elastase (PPE), *Cela1^−/−^* and WT mice had a similar level of emphysema. However, at 42 and 84 days, WT mice experienced progression of emphysema as evidenced by greater MLI values, while *Cela1^−/−^* mice did not. ANOVA p<0.001, Tukey *post hoc* comparisons are shown. Dashed line represents average MLI of WT PBS-treated mice. Representative 42-day photomicrographs are shown. Scale bar = 100 µm. (C) *Cela1^−/−^* mice exposed to 4 hours of cigarette smoke 5 days per week for 8 months had almost no detectable airspace simplification while such emphysema was detected in WT mice. ANOVA p-value < 0.0001, Tukey *post hoc* comparisons are shown. Representative tile scanned images of middle lobes are shown. Scale bar = 2,500 µm. (D) In comparing MLI of untreated mice aged 72 to 75 weeks, the MLI of wildtype mice was significantly increased while that of *Cela1^−/−^* was non-significantly larger (NS = not significant) and aged *Cela1^−/−^* had less age-related alveolar simplification than wildtype. ANOVA p-value < 0.001, Tukey *post hoc* comparisons are shown. Photomicrographs of Hart-stained lung sections of aged mouse lung show both preservation of alveolar structure in *Cela1^−/−^* mice and preservation of alveolar septal tip elastin bands (arrows). (E) By Western blot, the lungs of Cela1-/- mice have a greater amount of total lung tropoelastin (TE). Normalization is by total protein (TP) and comparison is by Welch’s t-test. *p<0.05, **p<0.01, ***p<0.001, ****p<0.0001

**Figure 5:**
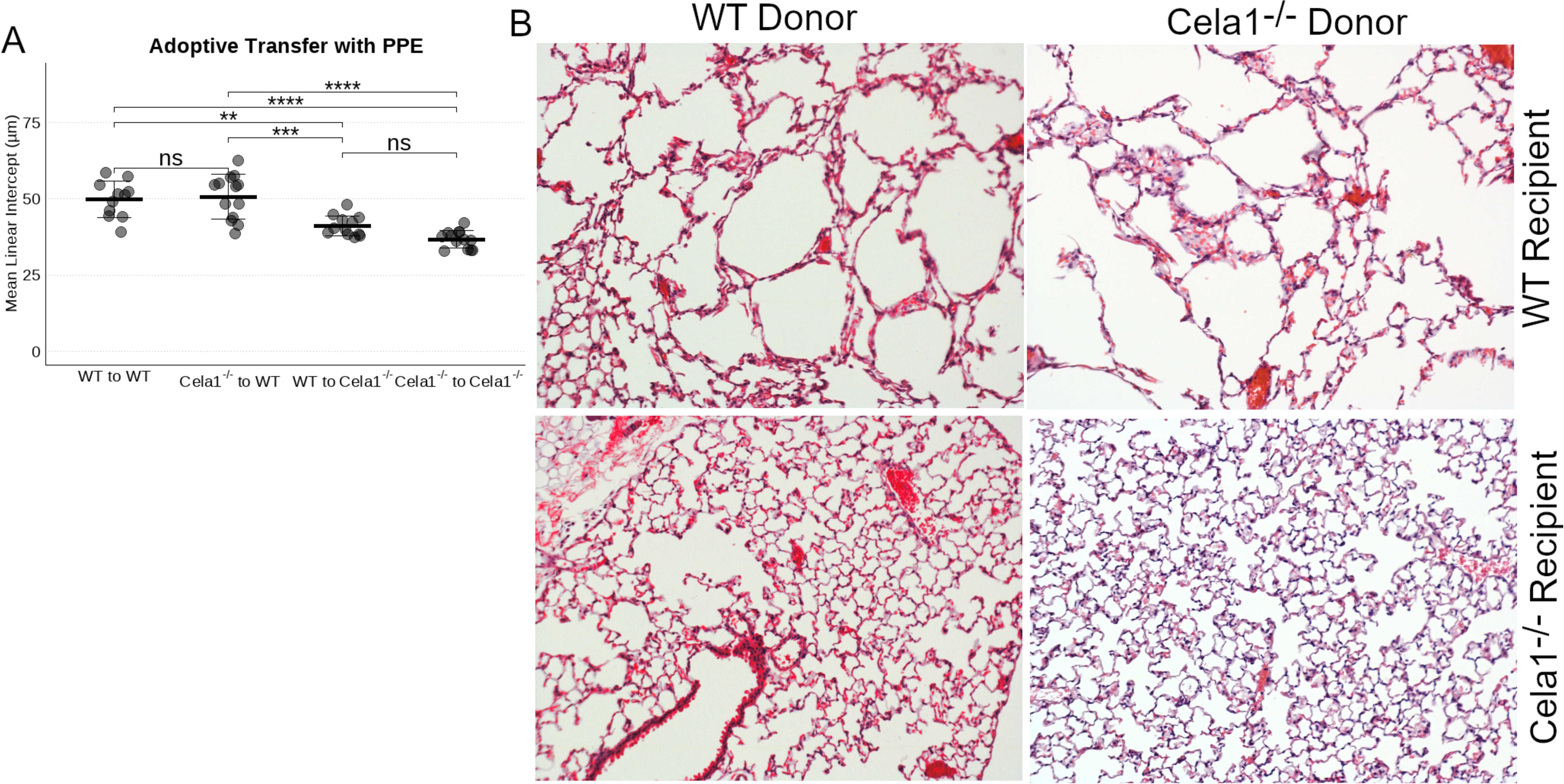
Bone Marrow Transplantation Does Not Impact CELA1-mediated Lung Remodeling. (A) Adoptive transfer of wildtype (WT) bone marrow to WT (n=11), *Cela1^−/−^* marrow to WT (n=13), WT to *Cela1^−/−^* (n=12), or *Cela1^−/−^*to *Cela1^−/−^*(n=13) with subsequent PPE-treatment showed airspace enlargement only in WT recipients. ANOVA p<0.0001. Comparisons by Tukey *post hoc* test. (B) Representative 10X photomicrographs. **p<0.01, ****p<0.00001

**Figure 6:**
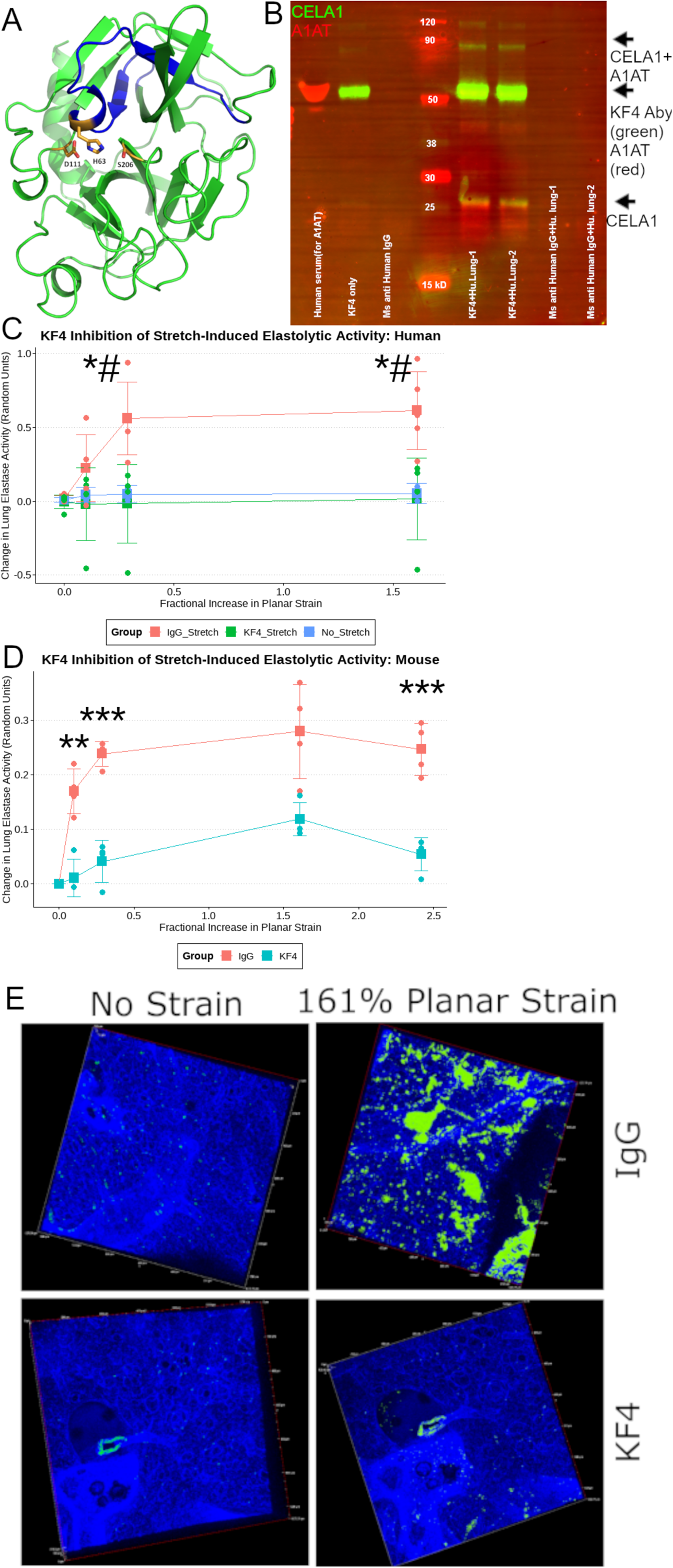
KF4 Anti-CELA1 Antibody. (A) The 3D model of human Chymotrypsin-like Elastase 1 retrieved from the Swiss-Model repository (Ref; ID: Q9UNI1) with the epitope detected by the KF4 antibody highlighted in blue. The three amino acids of the catalytic triad (H63, D111, and S206) are shown. The histidine of the CELA1 catalytic triad is within KF4 epitope. (B) Western blot of a human lung immunoprecipitation (IP) experiment using two human organ donor lung specimens. Lanes 1 and 2 are elutes from IP with KF4. Lanes 3&4 are elutes from IP with a mouse anti-human IgG antibody. Lanes 5&6 are elutes from IP with a polyclonal guinea pig anti-CELA1 antibody. For immunostaining, KF4 and anti-human α1-antitrypsin (AAT) antibodies are used as primary antibodies and detected with anti-mouse (green) and anti-rabbit (red) secondary antibodies respectively. The native 28 kDa CELA1 band is detected (green arrow) and a complex of CELA1 and AAT is detected at ~60 kDa (yellow arrow). (C) Five human lung sections were subjected to biaxial stretch in the presence of IgG or KF4 or incubated for equivalent times without stretch in the presence of elastin <u>in situ</u> zymography substrate. Stretch-inducible elastase activity was detected only in the IgG group. Comparisons at each time point are by ANOVA (p<0.05) with *post hoc* Tukey test p-values shown. *p<0.05 KF4 vs IgG, #p<0.05 KF4 vs. unstretched. (D) Stretch-inducible lung elastase activity in mouse lung sections showing ~80% reduction in activity at all points. Welch’s t-test comparison shown **p<0.01, ***p<0.001. (E) Representative images showing elastase activity in mouse lung sections subjected to biaxial stretch with IgG and KF4.

**Figure 7:**
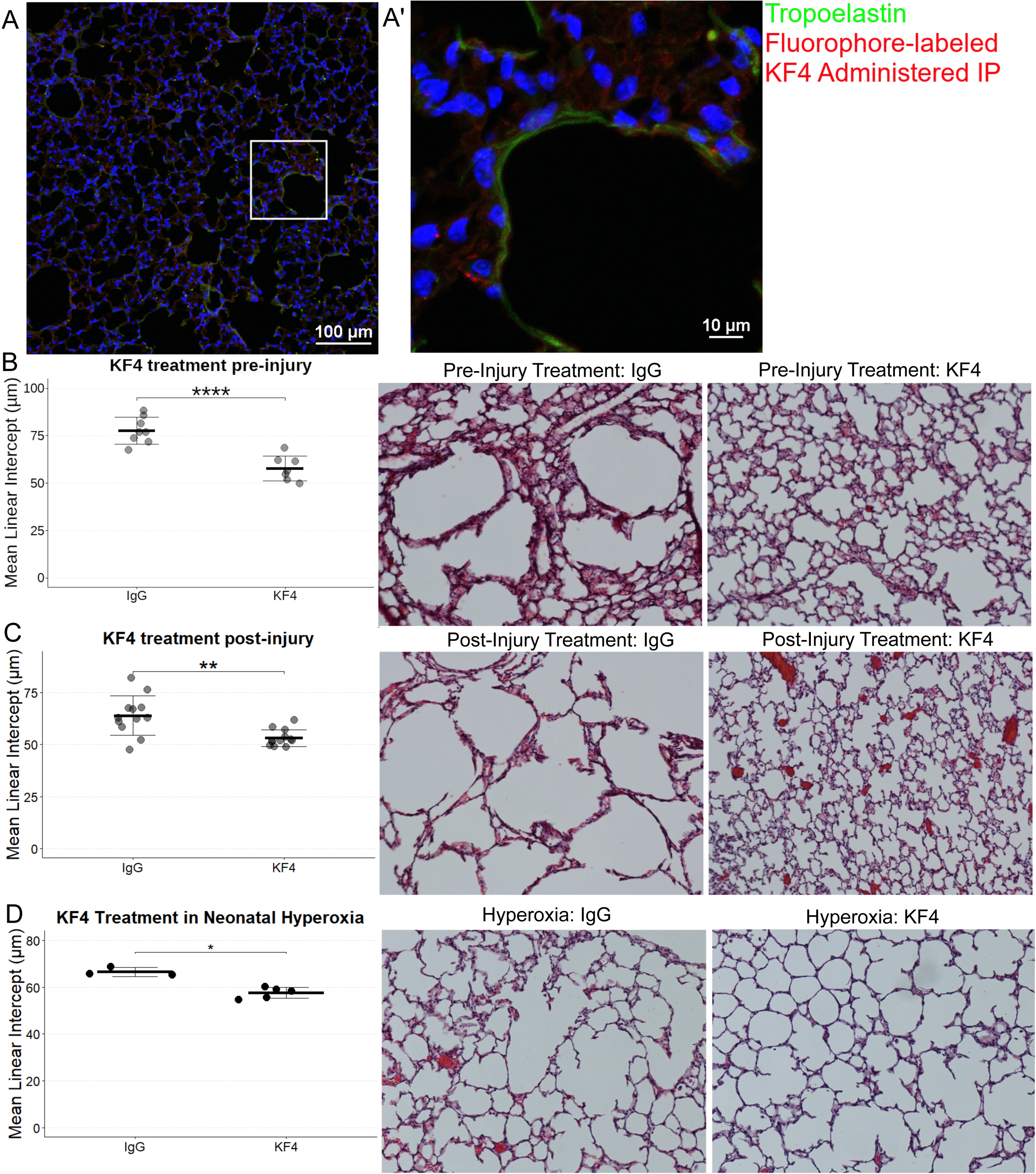
KF4 Treatment in Mouse Models of Alveolar Simplification. (A) Confocal image of a mouse that was administered fluorophore labeled KF4 (red) at a dose of 5 mg/kg 24 hours before tissue harvest. The lung section was counterstained for tropoelastin (green). KF4 signal is present throughout the interstitium. Scale bar = 100 µm. (A’) Magnified image of A showing a cell with increased binding of KF4, but also red signal throughout the lung matrix seemingly de-silhouetting cells suggested by central nuclei without red signal surrounding them. (B) Intraperitoneal administration of KF4 5 mg/kg once weekly at the time of treatment with tracheal PPE resulted in a significant improvement in airspace simplification. Comparison by Welch’s t-test. Representative 10X photomicrographs are shown. (C) Initiating the same KF4 therapy 7 days after PPE administration still resulted reduced airspace simplification. Comparison by Welch’s t-test. Representative 10X photomicrographs are shown. (D) Neonatal wildtype mice exposed to 80% oxygen for the first 14 days of life were treated with intraperitoneal KF4 on days 3, 7, and 10. This resulted in an over 50% reduction in airspace simplification. Representative 10X photomicrographs are shown. ANOVA p<0.0001, Selected Tukey post hoc comparisons are shown. **p<0.01, ****p<0.00001

### Bone Marrow Derived Immune Cells Do Not Mediate CELA1 Remodeling

To exclude the possibility that immune cells were playing a role in *Cela1*-mediated lung remodeling, we performed bone marrow transplantation experiments between wildtype and *Cela1^−/−^* mice. Irradiated wildtype or *Cela1^−/−^* mice were transplanted with WT or *Cela1^−/−^* bone marrow and then treated with PPE. WT recipients demonstrated an increase in mean linear intercept regardless of bone marrow genotype (Figure 5, Supplemental Figure 8), supporting AT2 cells as the relevant source of *Cela1* in airspace remodeling.

### Development and Validation of the KF4 Anti-CELA1 Antibody

We developed an anti-CELA1 monoclonal antibody by immunizing mice with four peptides corresponding to regions of human CELA1 with low mouse-human homology (Supplemental Table 1). Hybridomas were created, screened by ELISA and selected KF4 as our lead candidate based on inhibition of human lung elastase activity (Supplemental Figure 9). KF4 bound the N-terminal region of CELA1 in a region including the histidine of the catalytic triad (Figure 6A) (*24*). Immunoprecipitation of human lung homogenate with KF4 antibody identified both α1-antitrypsin complexed and native CELA1 (Figure 6B, Supplemental Figure 10). We then tested whether KF4 reduced stretch-inducible lung elastase activity. In organ donor lung sections, KF4 reduced stretch-inducible lung elastase activity to levels seen in unstretched lung (Figure 6C). In fresh mouse lung sections, KF4 reduced stretch-inducible lung elastase activity by 80% (Figure 6D-E). While the levels of strain in these *ex vivo* assays exceed those seen physiologically, they provide proof of principle that antibody neutralization of CELA1 inhibits strain-induced elastase activity.

### KF4 Penetrates into Lung Tissue Prevents Emphysema in Multiple Mouse Models

To identify the dose of KF4 needed to reduce mouse lung elastolytic activity, we performed dosing studies in PPE-treated mice and identified an effective intraperitoneal dose of 5 mg/kg once weekly (Supplemental Figure 11A). Using fluorophore-labeled KF4, we demonstrated penetration of KF4 into mouse lung tissue in a dose-dependent manner (Figure 7A, Supplemental Figure 11B). Since *Cela1^−/−^* mice demonstrated late protection against emphysema in the PPE model, we compared emphysema in IgG and KF4-treatment beginning at the time of injury and 7 days post-injury. The KF4 antibody prevented emphysema following tracheal PPE administration with both treatment strategies (Figure 7B-C). To test the efficacy of KF4 in a different model, we utilized the neonatal mouse hyperoxia model. When KF4 was administered at 3, 7, and 10 days of life, it also improved alveolarization compared to IgG (Figure 7D). Thus, systemically administered anti-CELA1 KF4 antibody penetrated lung tissue, inhibited stretch-inducible lung elastase activity, and preserved airspace architecture in multiple models of alveolar simplification.

## Discussion

We identified a unique role for *CELA1* in the strain-dependent remodeling of the peripheral lung and developed a monoclonal antibody that protects against strain-induced alveolar simplification. *CELA1*-mediated remodeling does not dependent upon inflammation or non-resident cells, but rather, *CELA1* is expressed by AT2 cells and is present in the lung extracelluar matrix but is only active with localized strain on that matrix. As such, it provides a mechanistic link between the protease-antiprotease model of emphysema and the structure-function one.

The biology of CELA1 is distinct from that of other serine and matrix metalloproteinases that have been implicated in airspace simplification. First, *CELA1* is expressed by a subset of AT2 cells while other serine proteases and matrix metalloproteinases are derived from immune cells. The supposition that *CELA1*-mediated lung matrix remodeling represents a different program than these myeloid-derived proteases is supported by (1) our analysis of human lung showing no correlation of *CELA1* mRNA levels with other proteases (except MMP12), (2) *CELA1* mRNA levels but not mRNAs of other proteases being correlated with lung elastase activity, and (3) a different pattern of emphysema protection than seen in other protease-focused emphysema studies. The effects of *neutrophil elastase*, *cathepsin G*, and *MMP12* showed protection at 21 days or less after injury (*25–28*). We found that protection of *Cela1^−/−^* mice beyond this time point. Consistent with our correlative analysis, others have found no correlation of MMP levels in bronchoalveolar lavage fluid and plasma with emphysema progression (*29*); although, sputum MMP12 levels are elevated in COPD (*30*). Alveolar macrophage depletion protected mice against elastase-induced emphysema at 7, 14, and 21 days (*31*), and much work has shown the interaction of MMPs and immune cells in emphysema (*32*). Similar to emphysema models, in the mouse hyperoxia model of BPD, MMPs, neutrophil elastase, and immune cell activation have all been shown important (*33–35*). Most work in age-related airspace simplification has focused on senescence and altered inflammatory responses (*36*). Although in a sense this work on *CELA1* falls into the 50-year-old protease-antiprotease paradigm, the novelty of *CELA1* lies in the cell type in which it is expressed, in its being only recently described, and the biomechanical mechanism by which it works.

*CELA1* mechanistically links the protease-antiprotease and the biomechanical models of emphysema. CELA1 binds and cleaves non-crosslinked hydrophobic domains of elastin (*6*), and these domains are only exposed with strain. Our findings that (1) CELA1 binding to human lung elastin is enhanced by strain, (2) CELA1 inhibition reduces stretch-induced lung elastolytic activity, (3) *Cela1^−/−^* mice are protected from late but not initial airspace simplification, and (4) that post-injury treatment with KF4 is just as efficacious as pre-injury treatment all support CELA1 as being a key mediator in localized strain-mediated airspace simplification. In this model, CELA1 is expressed by AT2 cells and binds and cleaves tropoelastin with strain. This increases strain in adjacent fibers which in turn exposes additional proteolytic sites (Figure 8A-C). KF4 binds the CELA1 catalytic domain and prevents elastolysis (Figure 8D). At the microstructural level, alveolar walls can be modeled as thin, geometrically shaped membranes under low strain suspended by thin elastic fibers—the fibers in the alveolar wall. These thin fibers are in turn supported by the more rigid fibers of the alveolar duct openings which are interconnected and supported by progressively larger caliber and less compliant fibers of the small and large conducting airways and vessels and to the pleural surface. Strain is distributed over the entire network. Failure of one alveolar wall necessitates increased strain on adjacent walls which both distorts local architecture and causes an imbalance of strain on additional alveolar walls. Each wall has a threshold beyond which it too will fail imparting additional strain in its neighbors predisposing them to failure (*11, 13*) (Figure 8E). The biomechanical model has been validated experimentally by showing that emphysema develops in regions of mouse lung not directly exposed to PPE (*37*). Furthermore, the role of *CELA1* in age-associated alveolar simplification is consistent with the accelerated aging model of COPD. If the same mechanisms in Figure 8 are operative in the aging lung, then *CELA1* provides a mechanistic link between three models: protease/antiprotease model, biomechanical, and accelerated aging.

**Figure 8:**
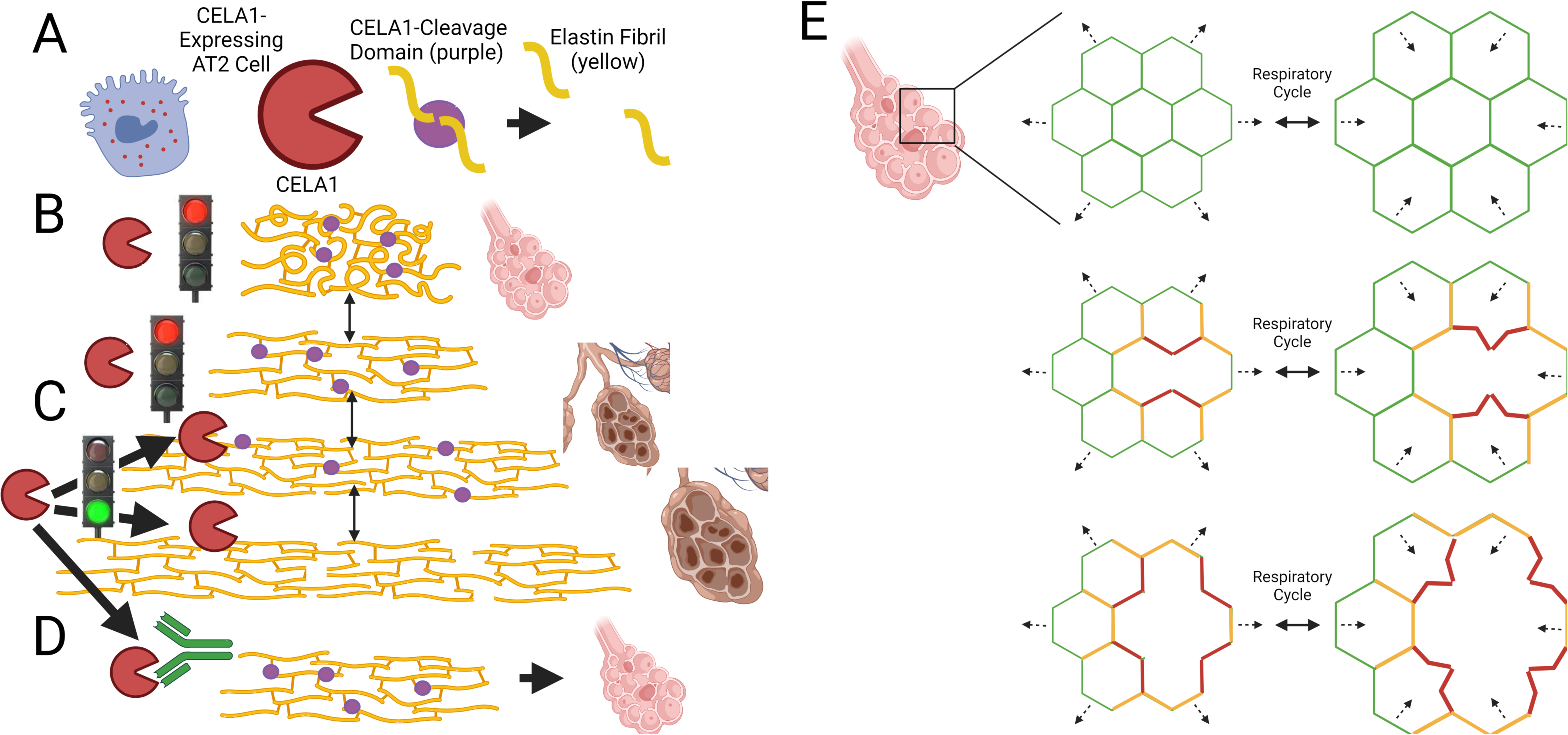
Mechanistic Model of CELA1 in Airspace Simplification. (A) CELA1 (red) is expressed in AT2 cells. It cleaves the hydrophobic domains of tropoelastin (purple circles). (B) Under normal conditions, throughout the respiratory cycle, CELA1 proteolytic sites are not accessible. (C) Under conditions of increased strain, CELA1 proteolytic sites are accessible and CELA1 cleaves those elastin fibers. This leads to increased strain on adjacent fibers and more cleavage. (D) The KF4 antibody binds the CELA1 catalytic triad, prevents strain-induced remodeling, and preserves airspace architecture. (E) At the alveolar level, alveolar walls are modeled as a repeating geometric pattern. Under normal conditions, the strain of the respiratory cycle (dashed arrows) is evenly distributed (green = low strain). After destruction of an alveolar wall, there is a high level of strain in adjacent alveolar walls (red) and moderate strain one generation away (orange). High strain walls are deformed with respiration leading to CELA1-mediated elastolysis and destruction. This leads to a cycle of progressive airspace destruction and emphysema. This cycle can be arrested with KF4.

Our study is not without limitations. Batch-to-batch variation in PPE injury precluded comparison between experiments. The biaxial strain model applies levels of strain that would not be experienced in the human lung. While we cannot definitively state that the same process would occur under physiological strain levels, neither can we perform this experiment over the months to years over which emphysema progresses. We cannot rule out the possibility that some absent role of *Cela1* on mouse lung development imparts resistance to postnatal lung injury; however, the protection conferred by anti-CELA1 KF4 antibody in multiple models makes this less likely. Lastly, we did not rigorously investigate whether *CELA1* could be upstream of other alveolar simplification programs, and it is possible that our *in vivo* effects are not direct ones.

## Conclusions

The strain-dependent elastolytic activity of CELA1 is an important factor in airspace simplification at all stages of postnatal lung life, and neutralizing CELA1 with the KF4 antibody represents a potential therapy for BPD, COPD, and other disorders of progressive airspace simplification.

## Materials and Methods

### Animal Use

Animal use was approved by the CCHMC Institutional Animal Use and Care Committee (2020-0054). Except for antibody production, all mice were on the C57BL/6 background, obtained from Jackson Laboratories, and equal proportions of male and female mice used in all experiments. Mice were maintained in a barrier facility with access to food and water *ad libidum* and 12-hour light-dark cycles.

### Hyperoxia Model

Paired female mice were bred and on PND1 pups were equally distributed between the two dams to give eight mice per litter. Pups were exposed to room air or hyperoxia with switching of dams between conditions every two days. On PND14, pups were either sacrificed or moved to room air with the same dam for 14 days and then sacrificed.

### Porcine Pancreatic Elastase Model

Eight- to ten-week-old mice were anesthetized with 2% isoflurane and suspended on an intubating board by the front incisors. Two units of porcine pancreatic elastase (Sigma, E1250) in 100 µL PBS was placed in the posterior oropharynx and aspirated by the breathing mouse.

### Cigarette Smoke Model

Beginning at 10-12 weeks of age, mice were exposed to 4 hours of cigarette smoke five days per week for eight months using a Teague TE-10z smoking machine using smoke generated from 3R4F Kentucky Reference Cigarettes (University of Kentucky) at a concentration of 150 mg/m^3^ total suspended particulates.

### Partial Pneumonectomy Model

Using previously described methods (*38*), the left lung of an anesthetized mouse was removed, mouse recovered, and tissues collected at 7 and 28 days.

### Adoptive Transfer

Eight- to 10-week-old mice had ablation of bone marrow with *** for *** and rescued with tail vein injection of *** cells from the bone marrow of donor mice. Mice were allowed to recover for 4 weeks prior to additional experimentation.

### Aging Model

Mice were sacrificed and collected at 72 to 75 weeks of age without any intervention.

### Mouse Lung Tissue

#### Tissue Processing and Morphometry

After anesthetization and sacrifice by exsanguination, the mouse thorax was opened, left bronchus ligated, left lung snap frozen, trachea canulated, right inflated with at 30 cm H_2_O pressure in 4% PBS, fixed overnight, lung lobes separated, and paraffinized. Lung lobes were randomly oriented and embedded, sectioned, and stained using hematoxylin and eosin. Using the methods of Dunnill (*39*), mean linear intercepts were determined in five field per lobe. Left lungs were homogenized with extraction of RNA using RNEasy columns or processed for protein analysis.

### Human Lung Tissue

Human tissue utilized under a waiver from the CCHMC IRB (2016-9641). COPD lung specimens were obtained from the NHLBI Lung Tissue Consortium, control and COPD specimens from the National Jewish Health Human Lung Tissue Consortium and from The Ohio State University Human Lung Tissue Consortium.

### Enzymatic Assays

Human lung protease, gelatinase, and elastase activity was quantified using Enzcheck fluorometric assays (ThermoFisher) and elastase activity defined as rate of signal change. For comparative inhibition assays (i.e. antibody inhibition) the fractional change vs. untreated was used for comparisons.

### *Ex vivo* Lung Stretch

Using a previously described lung stretching technique and device (*6, 7, 40*), the stretch-dependent binding of recombinant CELA1, albumin, and elastolytic activity of mouse and human lung sections was determined.

### Proximity Ligation *in situ* Hybridization (PLISH) and Immunofluorescence

Using oligos in Supplemental Table 2 and previously published methods (*41*), *CELA1* and *Sftpb* mRNAs were visualized with immunofluorescent co-staining using antibodies outlined in the supplement.

### PCR

Taqman and Sybr Green PCR was used for mouse and human lung PCR using primers in Supplemental Tables 3&4.

### anti-CELA1 Monoclonal Antibody Generation and Screening

CD-1 female mice were immunized with CELA1 peptides (Supplemental Table 1) conjugated to CRM197. Each mouse received a primary, subcutaneous immunization of 20 ug of the conjugate with 50% Titermax Gold adjuvant. The mice received subsequent immunizations of 20 ug (day 21) and 10 ug (day 35). Following the day 35 immunization serum samples were obtained and tested for reactivity with CELA1. Based on the results of the serum titration, one mouse was selected for hybridoma production. The mouse received an intravenous immunization of 2 ug of conjugate and three days later the mouse was euthanized and the spleen excised for hybridoma formation with SP2/0. Supernatants from the resulting hybridomas were tested for reactivity with CELA1 and were subsequently expanded and cloned. Antibodies from the cloned culture, derived from serum-free medium, was used in the studies reported here. Isotyping was performed using the Iso-Gold Rapid Mouse-Monoclonal Isotyping Kit (BioAssay Works Cat# KSOT03-010)

### Immunoprecipitation, Western Blot, and Proteomics

Ten mg of lung homogenates from two organ donors was incubated with 100 µg KF4 antibody and antibody precipitated using Protein A/G beads, washed, and eluting using loading dye. Western blots were performed using KF4 antibody, anti-Alpha 1 antitrypsin antibody (Abcam, ab133642), and anti-tropoelastin antibody (Abcam, ab21600). Revert total protein stain was used for normalization (LICOR, 926-11011). Silver stain was performed (Santa Cruz Biotechnology, sc-2003).

### Single Cell mRNA-Seq Analysis

Both mouse (*14*) and human (*15*) datasets were similarly analyzed to identify CELA1-expressing cells and perform gene set enrichment analysis of expressing vs non-expressing alveolar type 2 cells using Seurat(*42*) and enrichR (*43*) using scripts included in supplementary materials.

### Statistical Methods

Using R version 4.0.2 (*44*), the following packages were used for statistical comparisons and graphics generation: ggpubr (*45*), gridExtra (*46*), cowplot (*47*), ggplotify (*48*), corrplot (*49*), and rstatix (*50*). For parametric data, Welch’s t-test and ANOVA with Holm-Sidac post hoc test were used, and for non-parametric data, Wilcoxon rank-sum and Kruskall-Wallis with Dunn’s *post hoc* test were used. For correlative analyses, Pearson’s correlation with Bonferroni correction for multiple comparisons was used. Parametric data is displayed as line and whisker plots with center line representing mean and whiskers standard deviation. Non-parametric data is displayed as box plots with center line representing median value, boxes representing 25th to 75th percentile range, and lines representing 5th to 95th percentile range. For both plot types dots represent individual data points. For all analyses p-values of less than 0.05 were considered significant.

## Data Availability

After peer-review publication, the data in this manuscript will be made available upon request.

## Acknowledgements

Reagents and Tissues: We would like to acknowledge Dr. William Janssen and the National Jewish Health Human Lung Tissue Consortium in Denver, Colorado and Dr. Meghan Ballinger and the Ohio State University Lung Tissue Repository for providing control lung tissue specimens. This study utilized biological specimens and data provided by the Lung Tissue Research Consortium (LTRC) supported by the National Heart, Lung, and Blood Institute.

Core Services: Confocal imaging support was provided by the Cincinnati Children’s Hospital (CCHMC) Confocal Imaging Core. Adoptive transfer experiments were supported by the CCHMC Irradiator Core. Proteomics Support was provided by the University of Cincinnati Proteomics Laboratory.

## Funding

Innovation Fund, Innovation Ventures, Cincinnati Children’s Hospital (Varisco)

A1 Foundation Research Award 498262 (Varisco)

NIH/ NHBLI R01HL141229 (Varisco)

NIH/NHLBI K08HL131261 (Varisco)

NIH/NHLBI R01 HL147920 (Zemans)

NIH/NHLBI R01 HL131608 (Zemans)

## Competing Interests

Cincinnati Children’s Hospital and Dr. Varisco hold patents for KF4-based therapies for the treatment of AAT-deficient and non-AAT deficient emphysema (Case 2016-0706).

## Figure Legends

**Supplemental Figure 1:**
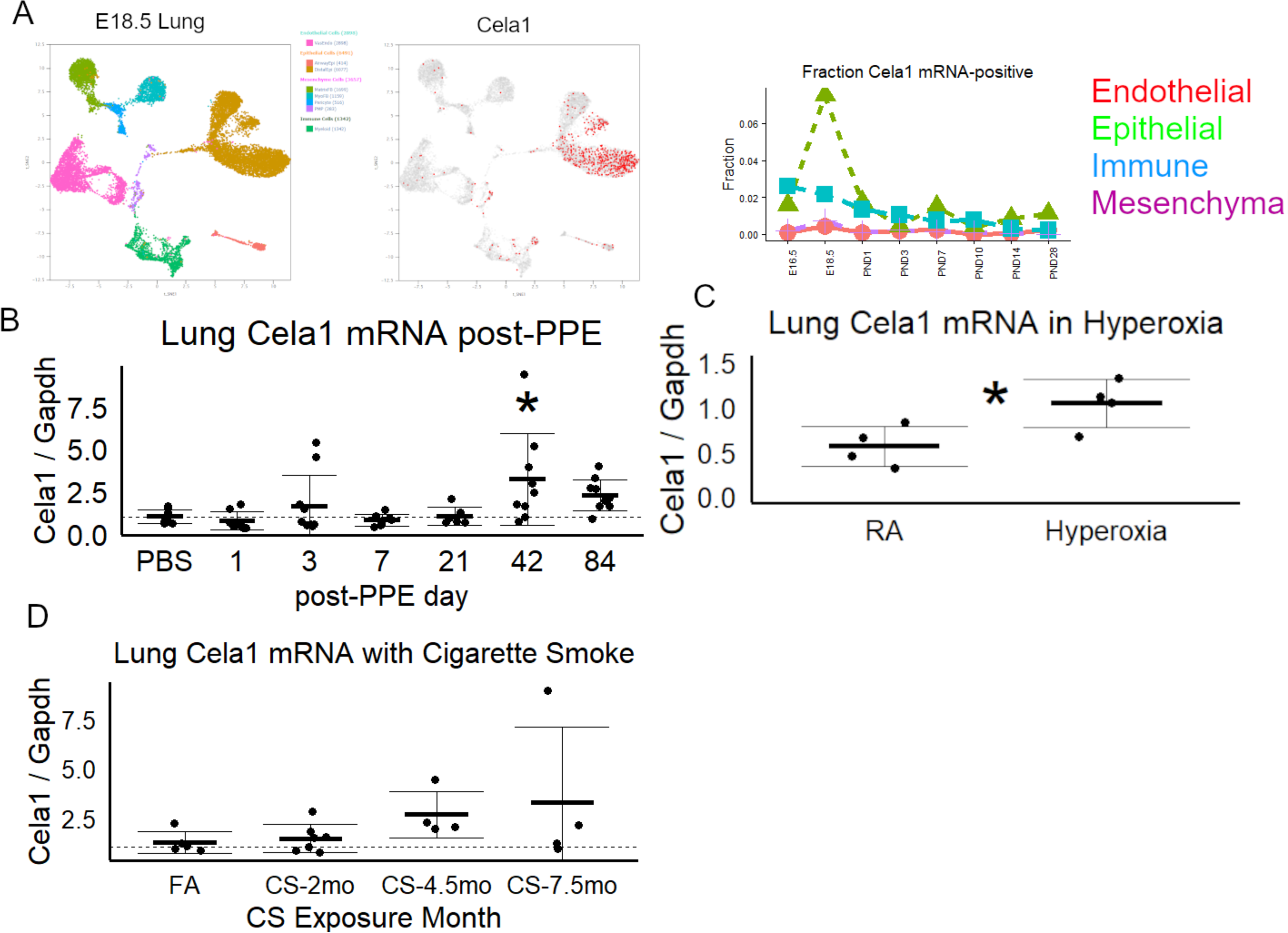
Lung Cela1 mRNA Levels in Development and Disease. (A) During lung development, there is an increase in *Cela1* mRNA-containing distal lug epithelial cells at E18.5 (data from LungGENS). (B) At sequential timepoints after airway administration of PPE, Cela1 mRNA levels were increased from baseline in some specimens with the increase at 42 days compared to PBS being significant by ANOVA with Dunnett *post hoc* test. (C) After 14 days of exposure to 80% oxygen, neonatal mouse lung *Cela1* mRNA levels were about twice that found at baseline as compared by Welch’s t-test. (D) Compared to filtered air, lung Cela1 mRNA levels tended to increase in mice exposed to cigarette smoke. *p<0.05, PPE=Porcine Pancreatic Elastase, CS=Cigarette Smoke, Cela1=Chymotrypsin-like Elastase 1 Gapdh = Glyceraldehyde-3-phosphate Dehydrogenase FA=Filtered Air

**Supplemental Figure 2:**
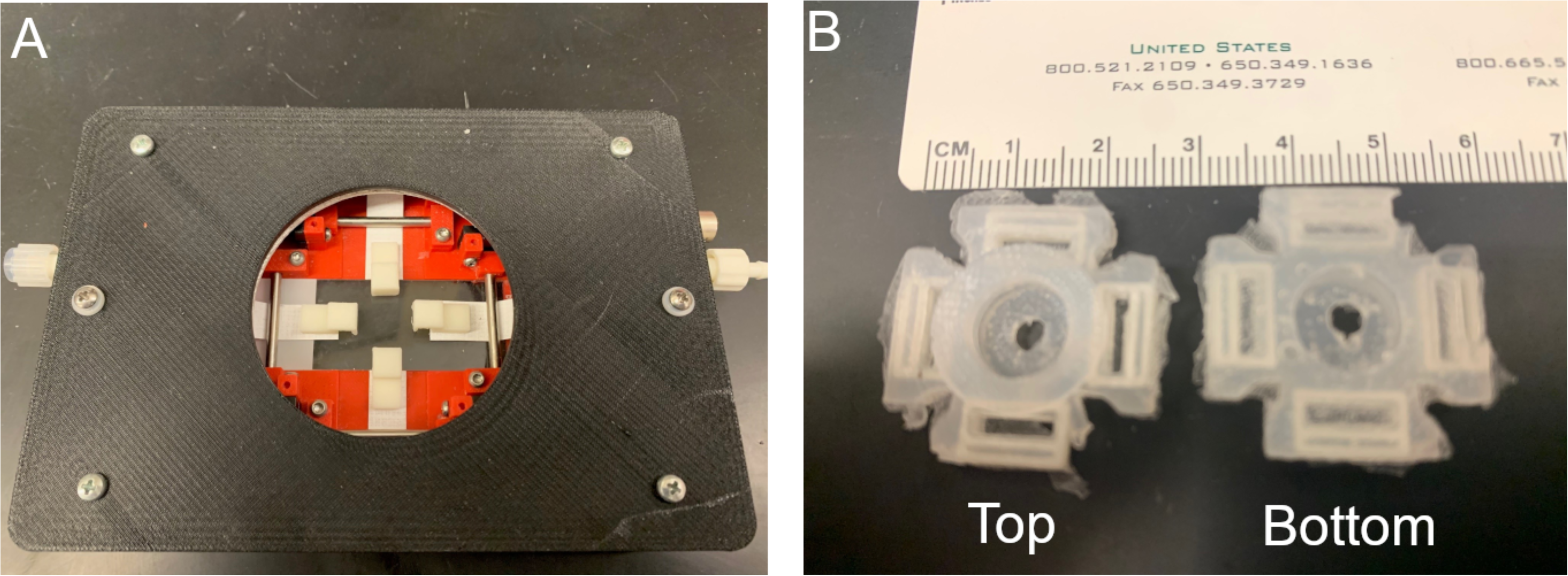
Biaxial Stretch of Human Lung Tissue. (A) Image of a 3D-printed, confocal microscope stretching device. The device fits into a K-mount of a Nikon A1 confocal microscope and contains 4 small motors which incrementally turn a Tyvek strip to which is attached printed clips. These clips are secured to a silicone mount. (B) Silicone mount which is created using 3D-printed molds with embedded eyelets to which the clips are secured. 100 µm sections of human lung are secured to the underside of the mount at four points using surgical glue.

**Supplemental Figure 3:**
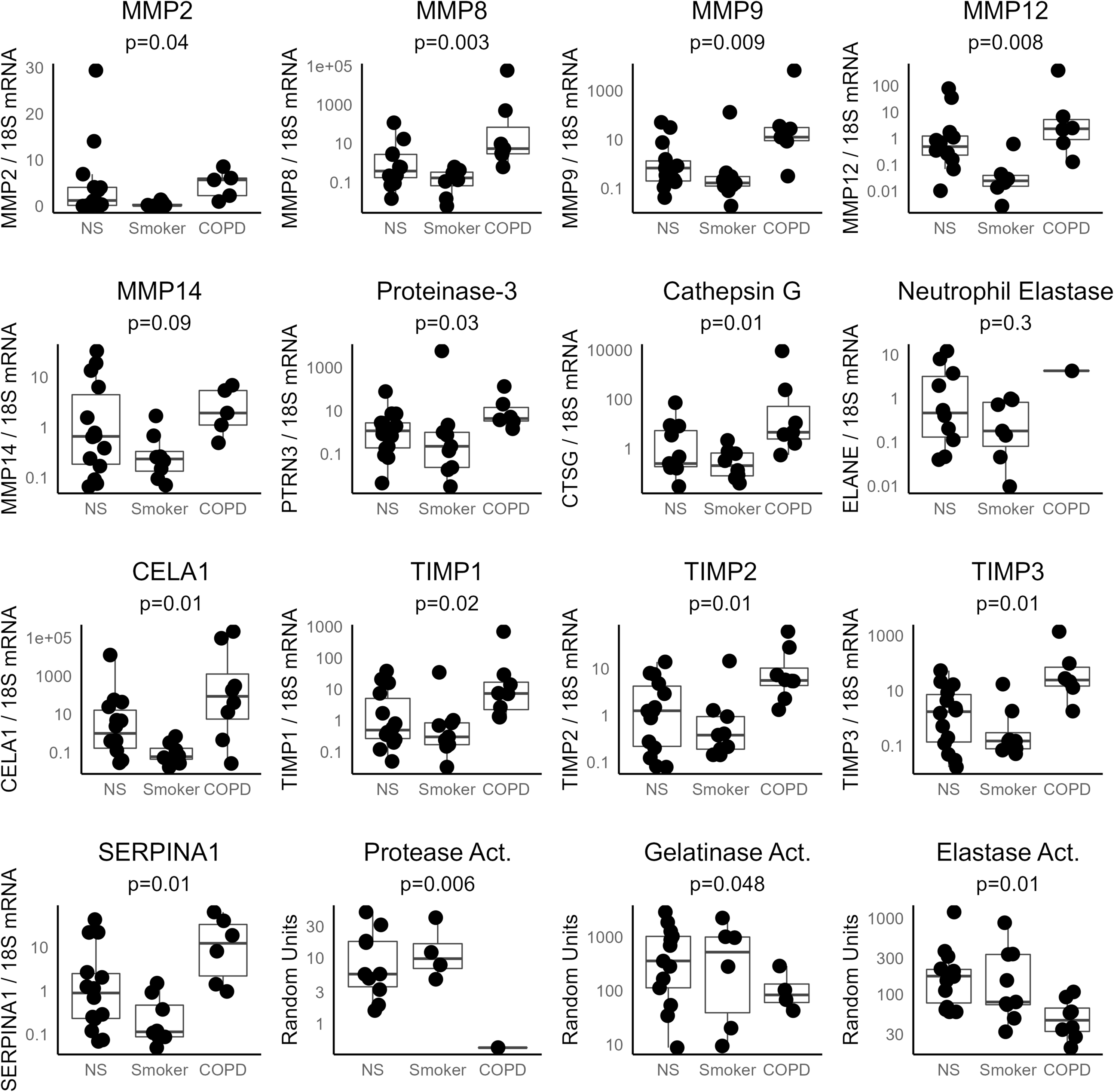
mRNA Levels of COPD-associated Proteases and Anti-Proteases. The mRNA levels of matrix metalloproteinase 2 (MMP2), MMP8, MMP9, MMP12, MMP14, Proteinase-3, Cathepsin G, Neutrophil Elastase, CELA1, Tissue Inhibitor of Metalloproteinase-1 (TIMP1), TIMP2, TIMP3, and α1-antitrypsin (SERPINA1) were generally higher in COPD than smoker controls and non-smoker (NS) controls. Tissue protease, gelatinase, and elastase activities were all lower in COPD however. Kruskal-Wallis p values are shown.

**Supplemental Figure 4:**
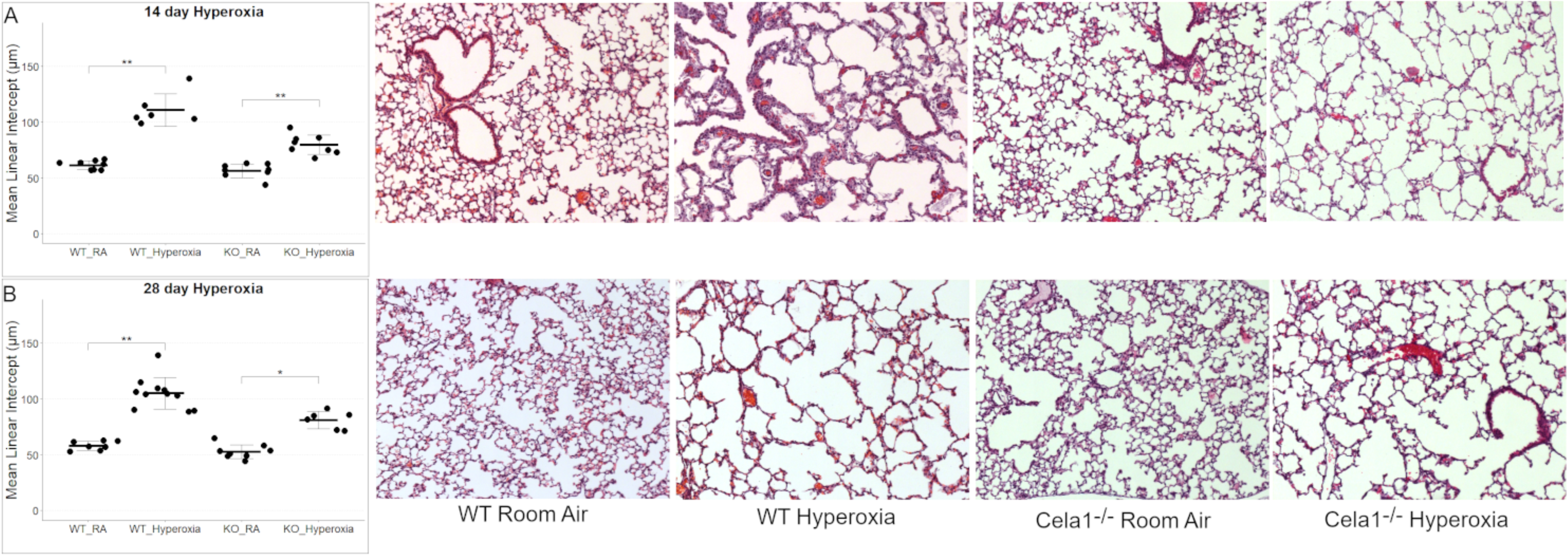
Protection of Cela1-/- Mice in Hyperoxia Model. (A) Mice were exposed to 80% oxygen from postnatal days 1 to 14. At 14 days, both *Cela1^−/−^* and WT mice had evidence of alveolar simplification, but the simplification was less in *Cela1^−/−^* mice. ANOVA p<0.01. Tukey *post hoc* comparisons are shown. Representative 10X photomicrographs are shown to the right. (B) Fourteen days after removal from hyperoxia, the differences in airspace size persisted. ANOVA p<0.01. Tukey *post hoc* comparisons are shown. *p<0.05, **p<0.01

**Supplemental Figure 5:**
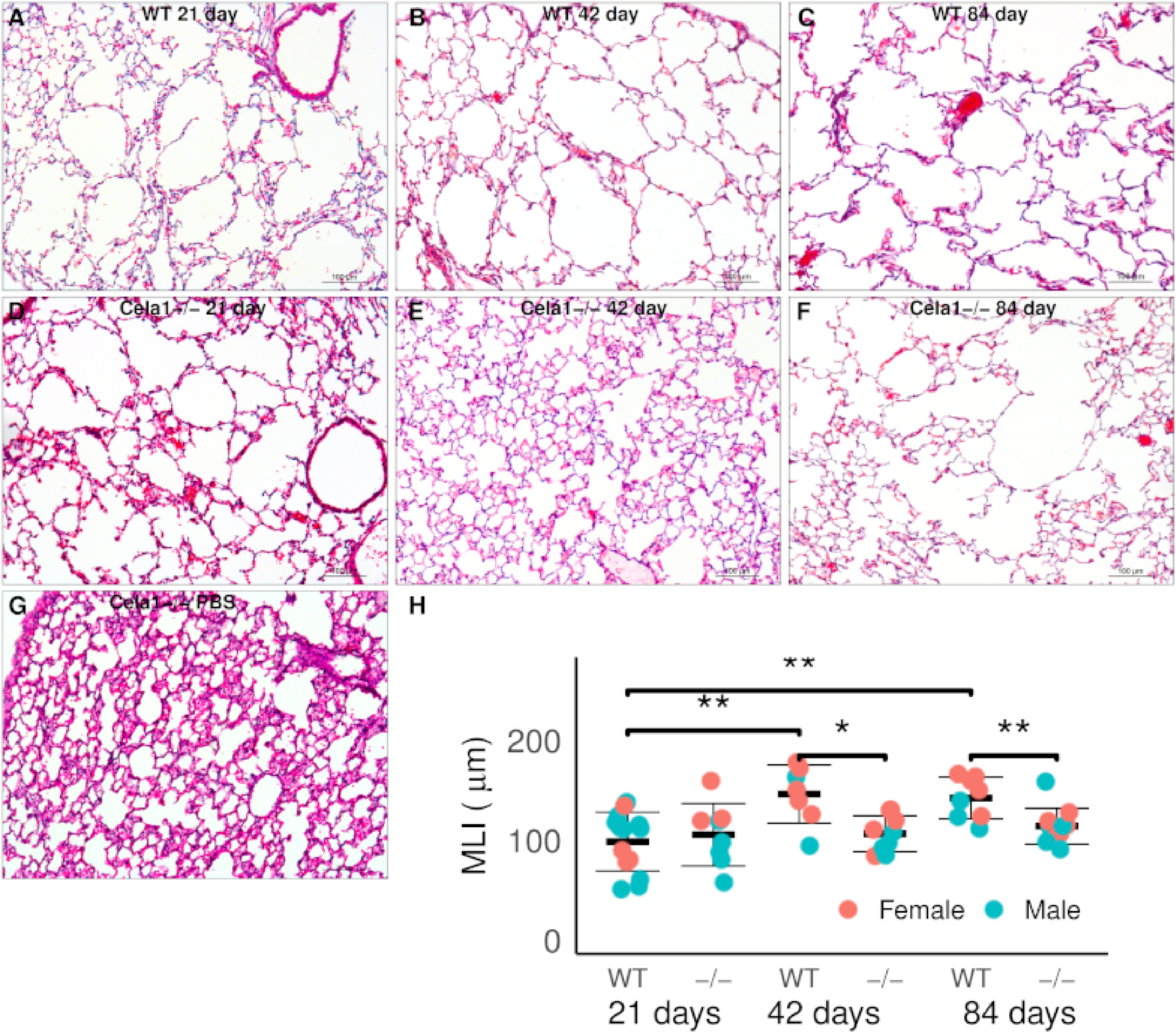
Protection of Cela1-/- Mice in PPE Model. (A) Mice were treated with 2 units of PPE instilled into the posterior nasopharynx. Treated wildtype (WT) mice had evidence of airway simplification at 21 days. 10X photomicrograph, scale bar = 100 µm. (B) This airspace simplification was worse at 42 days and (C) at 84 days. (D) *Cela1^−/−^* mice had a similar degree of airspace injury at 21 days. (E) However, at 42 and (F) 84 days, these mice did not experience progression of airspace simplification. (G) PBS-treated *Cela1^−/−^* mice had normal appearing airspace architecture. (H) Line and whisker plot showing the above differences. There was no difference in emphysema by sex. ANOVA p < 0.01. Tukey post hoc comparisons are shown. *p<0.05, **p<0.01

**Supplemental Figure 6:**
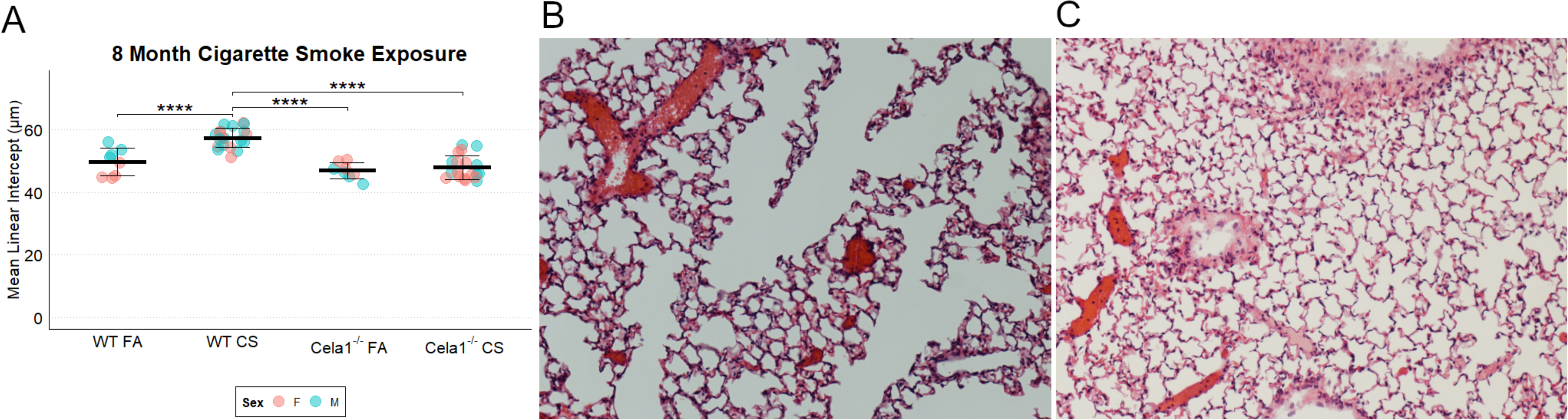
Protection of Cela1-/- Mice in Cigarette Smoke Model of Emphysema. (A) Compared to wildtype (WT) mice, *Cela1^−/−^* mice had no difference in airspace size at baseline but were protected against airspace simplification in response to cigarette smoke (CS) exposure. There was no impact of sex on airspace size. ANOVA p<0.00001. Tukey *post hoc* comparisons are shown. (B) Representative 10X photomicrograph of CS-exposed WT lung and (C) CS-exposed *Cela1^−/−^* lung.

**Supplemental Figure 7:**
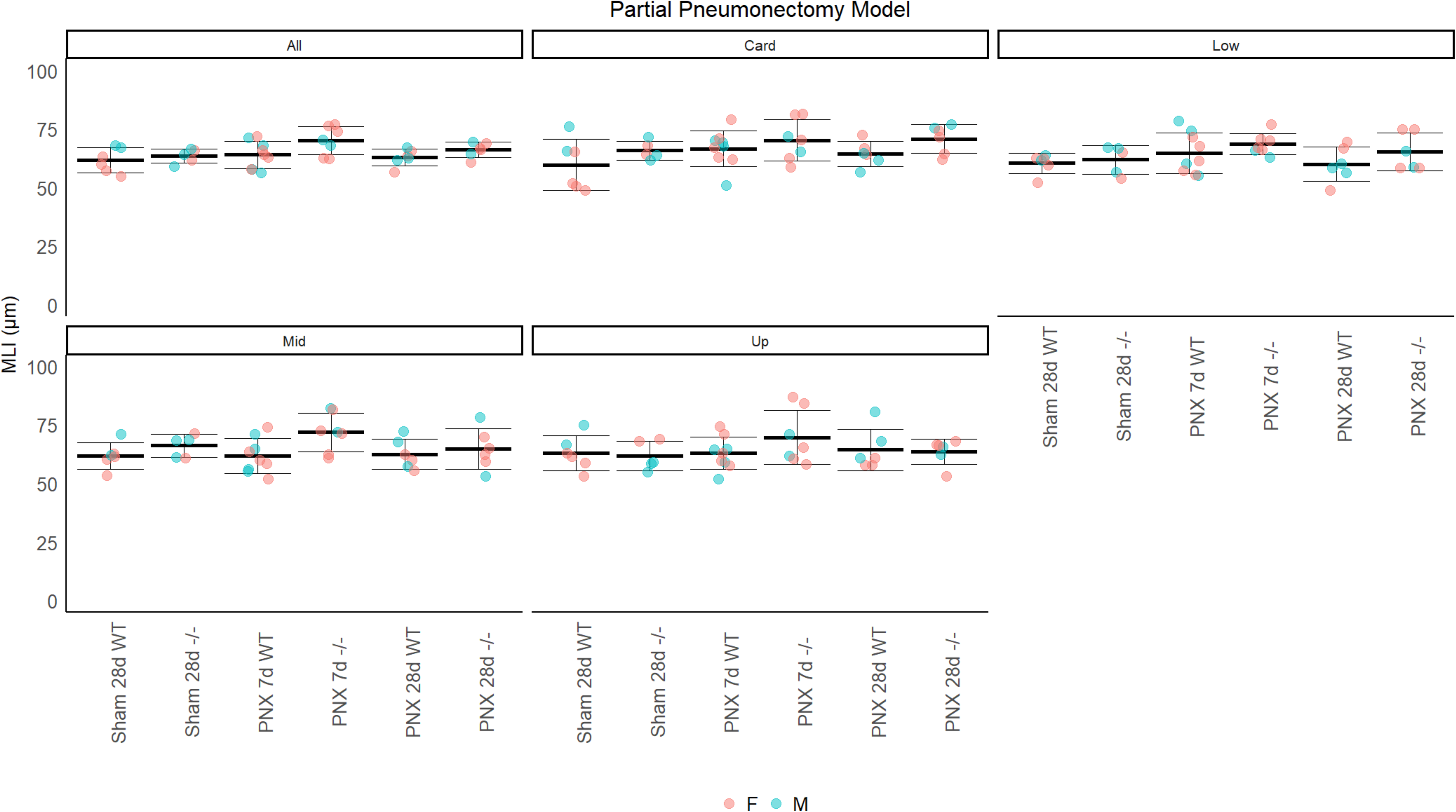
No Impact of CELA1 in Compensatory Lung Re-growth after Partial Pneumonectomy. (A) Wildtype (WT) and *Cela1^−/−^* (-/-) mice were subjected to left lung partial pneumonectomy, and the airspace size of the remaining lung lobes quantified at 7 and 28 days. Mean linear intercepts (MLI) were compared to Sham at 28 days. A small increase in MLI in *Cela1^−/−^* mice at 7 days was not significant and normalized by 28 days.

**Supplemental Figure 8:**
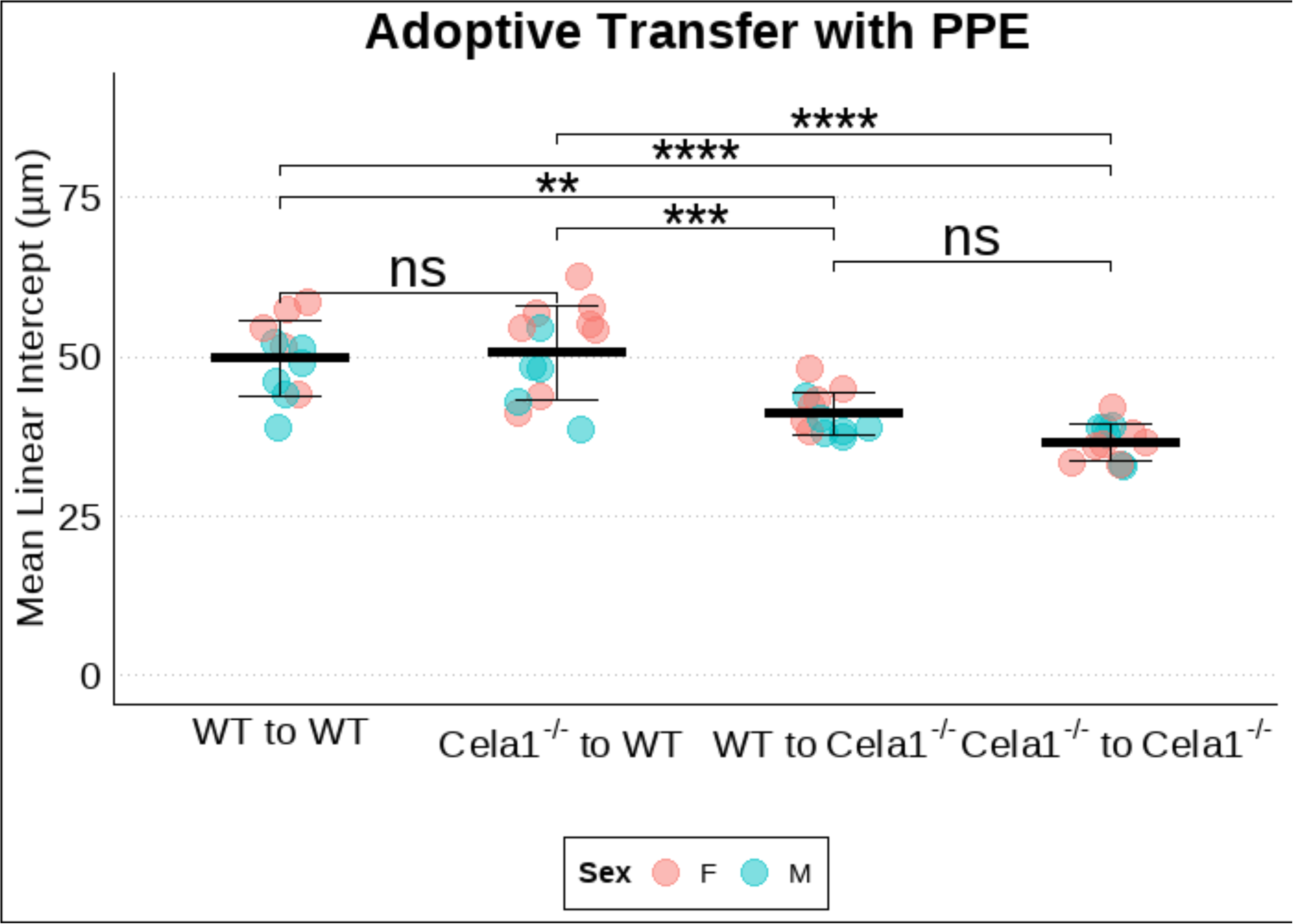
Protection of Cela1-/- Mice in PPE Model is Derived from Resident Lung Cells. (A) Adoptive transfer was performed between wildtype (WT) and *Cela1^−/−^* mice. After recover, mice were treated with airway porcine pancreatic elastase (PPE) and tissues collected at 42 days. *Cela1^−/−^* bone marrow did not confer protection to WT mice, and WT bone marrow did not increase sensitivity of *Cela1^−/−^* mice. ANOVA p<0.00001. Tukey post hoc comparisons are shown. **p<0.01, ***p<0.001, ****p<0.00001 ns=not significant

**Supplemental Figure 9:**
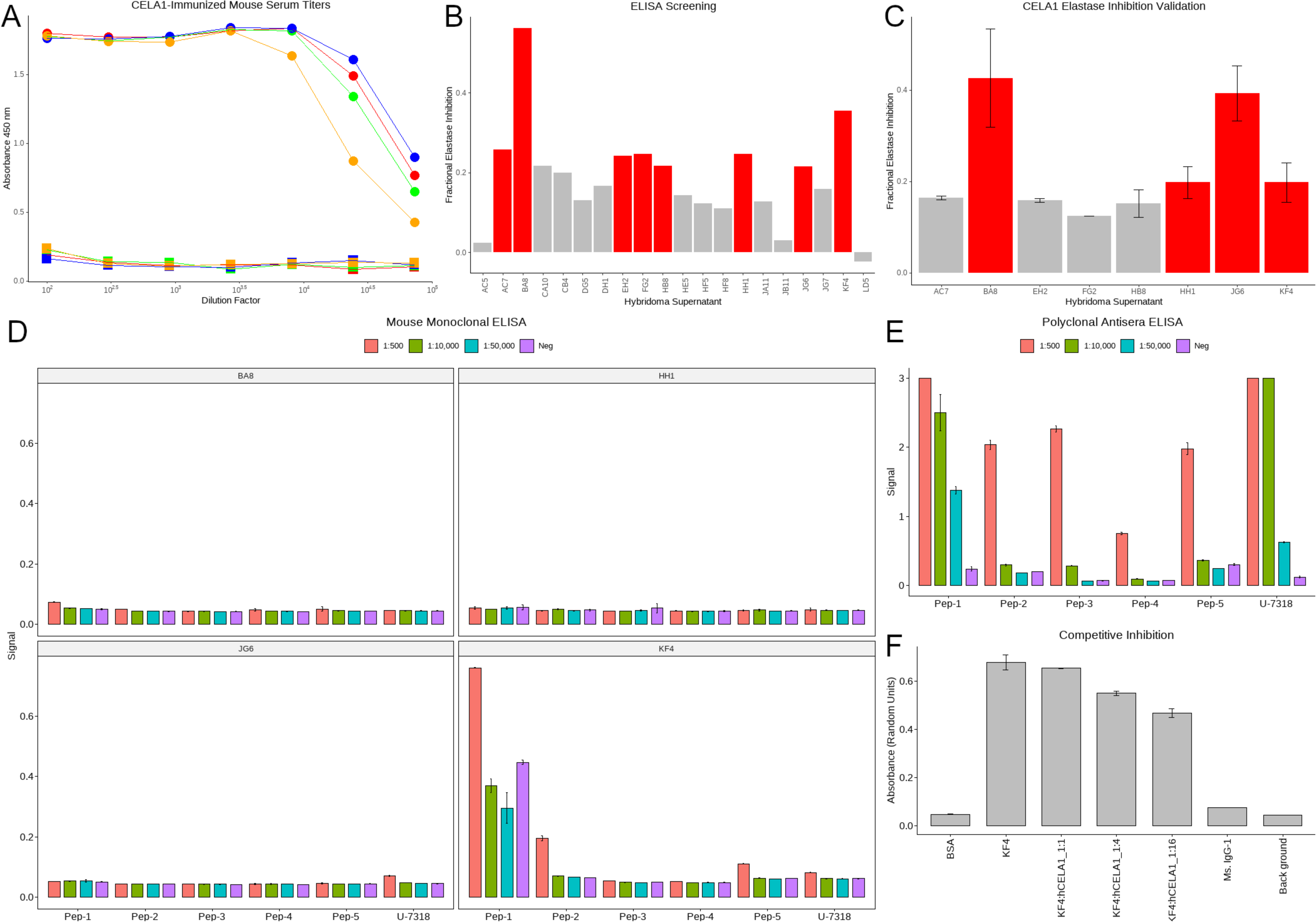
Identification of KF4 as Lead Candidate. (A) The serum of four mice immunized with human CELA1 peptides all demonstrated high titers in an ELISA against recombinant CELA1. (B) Hybridomas were created and screened and the eight clones with the highest titers selected for functional screening. (C) The four clones (BA8, HH1, JG6, and KF4) with the greatest inhibition of CELA1 elastolytic activity were selected. (D) The immunizing peptides were immobilized and used for ELISA. Only KF4 detected one of the peptides used for immunization. (E) As a positive control, an anti-CELA1 polyclonal antibody was used and detected all of the peptides. (F) KF4 was incubated with increasing ratios of recombinant CELA1 prior to testing by ELISA in a competitive inhibition assay. There was a serial reduction in ELISA signal.

**Supplemental Figure 10:**
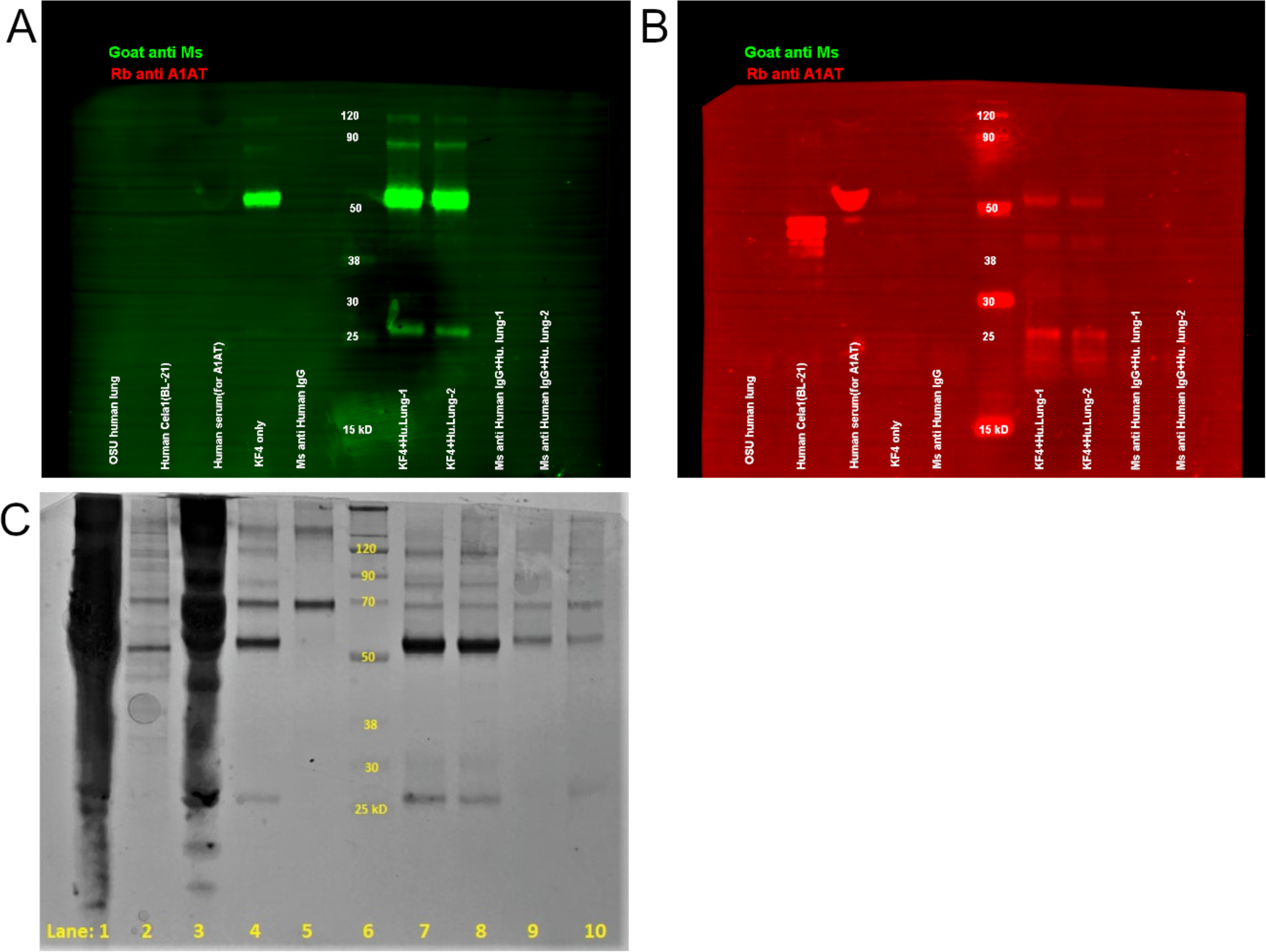
KF4 Dosing and Tissue Penetration. (A) Mice were treated with PPE, and at 42 days were treated with 2, 5, or 15 mg/kg KF4 or 15 mg/kg IgG. Lung elastase activation was reduced most prominently in the 5 mg/kg dose. n=4 per group. (B) KF4 was conjugated with AlexaFlour-647 and administered to mice at 0.2, 1, and 5 mg/kg. Tissues were collected, homogenized, and the fluorescence signal of 10 mg tissue quantified. Lung and serum Kruskal-Wallis p <0.05. Dunn *post hoc* test shown. *p<0.05, **p<0.01

**Supplemental Figure 11:**
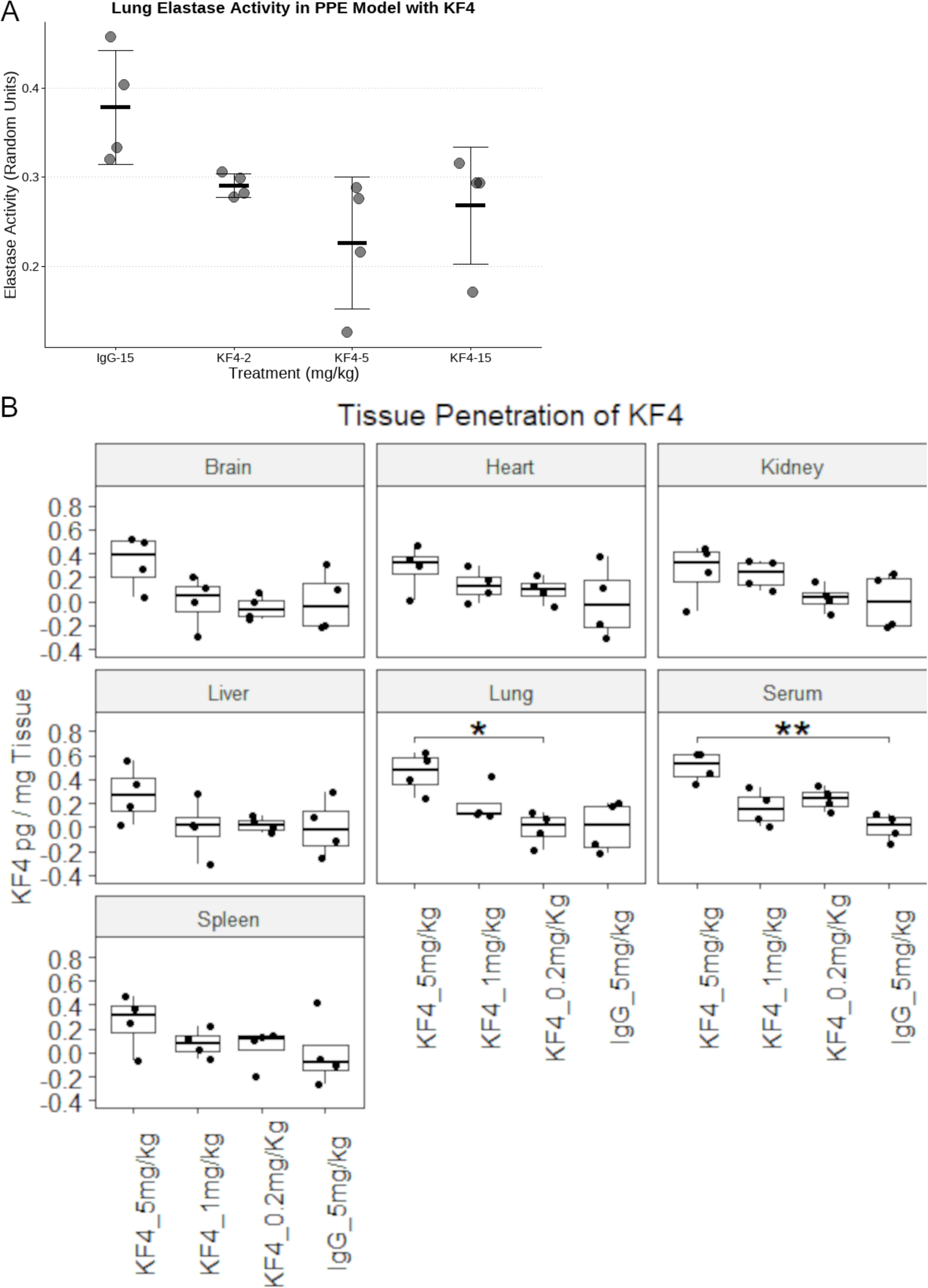

## Tables

**Supplemental Table 1:**
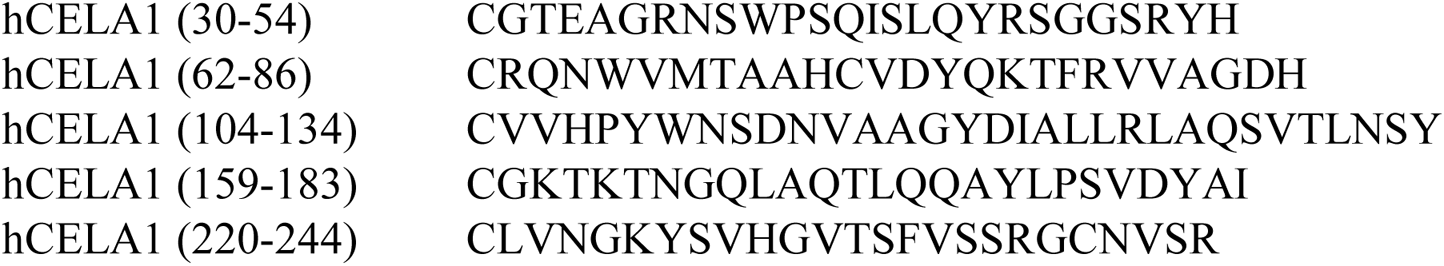
hCELA1 Peptides.

**Supplemental Table 2:**
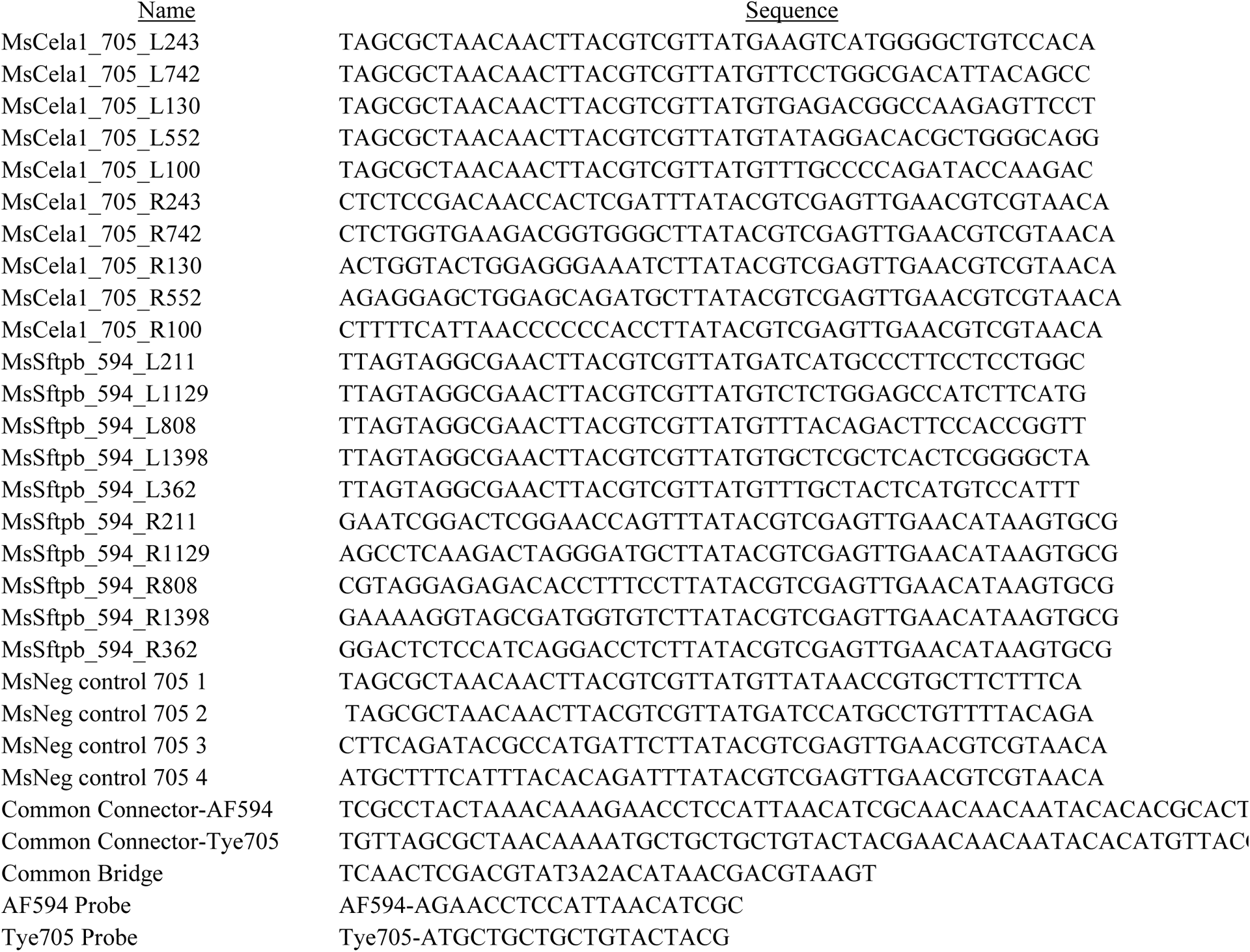
PLISH Primers, Bridges, and Probes.

**Supplemenal Table 3:** Taqman Primers.

| <u>Target</u> | <u>Primer Catalog Number</u> |
| --- | --- |
| Human Matrix Metaloproteinase-2 | 4331182_Hs01548727 |
| Human Matrix Metaloproteinase-8 | 4453320-Hs01029057 |
| Human Matrix Metaloproteinase-9 | 4453320-Hs00957562_m1 |
| Human Matrix Metaloproteinase-12 | 4448892-Hs00159178 |
| Human Matrix Metaloproteinase-14 | 4448892-Hs01037003 |
| Human Proteinase-3 | 4448892-Hs01553330_m1 |
| Human Neutrophil Elastase | 4331182-Hs00236952_m1 |
| Human Chymotrypsin-like Elastase 1 | 4331182Hs00608115_m1 |
| Human Cathepsin G | 4448892-Hs00175195_m1 |
| Eukaryotic 18S RNA | 4333760T |

**Supplemenal Table 4:**
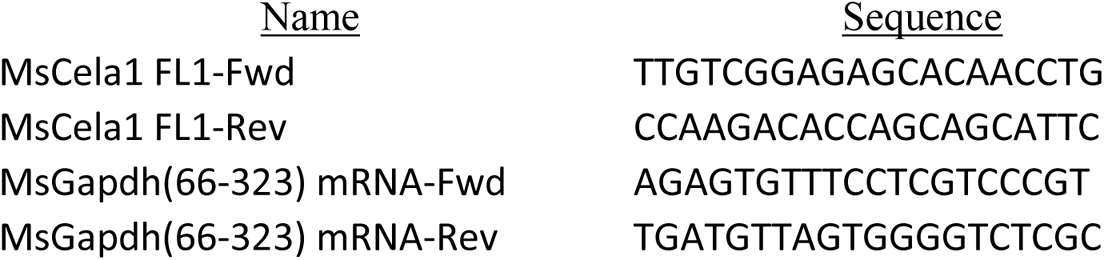
SybrGreen Primers.

## Notes

**Conflict of Interest Statement**: Brian Varisco and Cincinnati Children’s Hospital hold patents for the use of anti-CELA1 therapies and the KF4 antibody to prevent emphysema.

### Competing Interest Statement

Patents for use of anti-CELA1 therapies for alpha-1 antitrypsin-deficient emphysema and COPD.

### Funding Statement

Funding: A1 Foundation Research Award 498262 (Varisco), NHBLI R01141229 (Varisco), NHLBI K08HL131261 (Varisco).

### Author Declarations

Human tissue utilized under a waiver from the CCHMC IRB (2016-9641)

### Summary of Updates

Data on the use of KF4 anti-CELA1 antibody to prevent emphysema.

## References

1. S. M. May, J. T. C. Li, Burden of chronic obstructive pulmonary disease: Healthcare costs and beyond. Allergy Asthma Proc 36, 4–10 (2015).

2. B. R. Celli, J. A. Wedzicha, Update on Clinical Aspects of Chronic Obstructive Pulmonary Disease. New England Journal of Medicine 381, 1257–1266 (2019).

3. S. Y. Ash, R. San José Estépar, S. B. Fain, R. Tal-Singer, R. A. Stockley, L. H. Nordenmark, S. Rennard, M. K. Han, D. Merrill, S. M. Humphries, A. A. Diaz, S. E. Mason, F. N. Rahaghi, C. L. Pistenmaa, F. C. Sciurba, G. Vegas-Sánchez-Ferrero, D. A. Lynch, G. R. Washko, Relationship between Emphysema Progression at CT and Mortality in Ever-Smokers: Results from the COPDGene and ECLIPSE Cohorts. Radiology 299, 222–231 (2021).

4. J. Ozsvar, C. Yang, S. A. Cain, C. Baldock, A. Tarakanova, A. S. Weiss, Tropoelastin and Elastin Assembly. Frontiers in Bioengineering and Biotechnology 9 (2021) (available at https://www.frontiersin.org/article/10.3389/fbioe.2021.643110).

5. S. Liu, S. M. Young, B. M. Varisco, Dynamic expression of chymotrypsin-like elastase 1 over the course of murine lung development. American journal of physiology. Lung cellular and molecular physiology 306, L1104–16 (2014).

6. R. Joshi, A. Heinz, Q. Fan, S. Guo, B. Monia, C. E. H. Schmelzer, A. S. Weiss, M. Batie, H. Parameshwaran, B. M. Varisco, Role for Cela1 in Postnatal Lung Remodeling and AAT-deficient Emphysema. American journal of respiratory cell and molecular biology (2018), doi:10.1165/rcmb.2017-0361OC.

7. R. Joshi, S. Liu, M. D. Brown, S. M. Young, M. Batie, J. M. Kofron, Y. Xu, T. E. Weaver, K. Apsley, B. M. Varisco, Stretch regulates expression and binding of chymotrypsin-like elastase 1 in the postnatal lung. FASEB journal : official publication of the Federation of American Societies for Experimental Biology 30, 590–600 (2016).

8. C. B. Laurell, S. Eriksson, [HYPO-ALPHA-1-ANTITRYPSINEMIA]. Verh Dtsch Ges Inn Med 70, 537–539 (1964).

9. N. Mercado, K. Ito, P. J. Barnes, Accelerated ageing of the lung in COPD: new concepts. Thorax 70, 482–489 (2015).

10. H. Parameswaran, A. Majumdar, B. Suki, Linking microscopic spatial patterns of tissue destruction in emphysema to macroscopic decline in stiffness using a 3D computational model. PLoS Comput Biol 7, e1001125 (2011).

11. H. Hamakawa, E. Bartolak-Suki, H. Parameswaran, A. Majumdar, K. R. Lutchen, B. Suki, Structure-function relations in an elastase-induced mouse model of emphysema. Am J Respir Cell Mol Biol 45, 517–24 (2011).

12. S. Ito, E. P. Ingenito, K. K. Brewer, L. D. Black, H. Parameswaran, K. R. Lutchen, B. Suki, Mechanics, nonlinearity, and failure strength of lung tissue in a mouse model of emphysema: possible role of collagen remodeling. J Appl Physiol 98, 503–11 (2005).

13. B. Suki, S. Sato, H. Parameswaran, M. V. Szabari, A. Takahashi, E. Bartolak-Suki, Emphysema and mechanical stress-induced lung remodeling. Physiology (Bethesda, Md.) 28, 404–13 (2013).

14. J. Choi, J.-E. Park, G. Tsagkogeorga, M. Yanagita, B.-K. Koo, N. Han, J.-H. Lee, Inflammatory Signals Induce AT2 Cell-Derived Damage-Associated Transient Progenitors that Mediate Alveolar Regeneration. Cell Stem Cell (2020), doi:10.1016/j.stem.2020.06.020.

15. M. Sauler, J. E. McDonough, T. S. Adams, N. Kothapalli, J. S. Schupp, J. Nouws, M. Chioccioli, N. Omote, C. Cosme, S. Poli, E. A. Ayaub, S. G. Chu, K. H. Jensen, J. Gomez-Villalobos, C. J. Britto, M. S. Raredon, P. N. Timshel, N. Kaminski, I. O. Rosas, Single-cell RNA sequencing identifies aberrant transcriptional profiles of cellular populations and altered alveolar niche signalling networks in Chronic Obstructive Pulmonary Disease (COPD). medRxiv, 2020.09.13.20193417 (2020).

16. R. Jesudason, S. Sato, H. Parameswaran, A. D. Araujo, A. Majumdar, P. G. Allen, E. Bartolak-Suki, B. Suki, Mechanical forces regulate elastase activity and binding site availability in lung elastin. Biophysical journal 99, 3076–83 (2010).

17. P. Sakornsakolpat, M. McCormack, P. Bakke, A. Gulsvik, B. J. Make, J. D. Crapo, M. H. Cho, E. K. Silverman, Genome-Wide Association Analysis of Single Breath Diffusing Capacity of Carbon Monoxide (DLCO). https://doi.org/10.1165/rcmb.2018-0384OC (2019), doi:10.1165/rcmb.2018-0384OC.

18. P. Sakornsakolpat, J. D. Morrow, P. J. Castaldi, C. P. Hersh, Y. Bosse, E. K. Silverman, A. Manichaikul, M. H. Cho, Integrative genomics identifies new genes associated with severe COPD and emphysema. Respiratory research 19, 46 (2018).

19. 1000 Genomes Project Consortium, A. Auton, L. D. Brooks, R. M. Durbin, E. P. Garrison, H. M. Kang, J. O. Korbel, J. L. Marchini, S. McCarthy, G. A. McVean, G. R. Abecasis, A global reference for human genetic variation. Nature 526, 68–74 (2015).

20. Home - dbGaP - NCBI (available at https://www.ncbi.nlm.nih.gov/gap/).

21. M. J. Landrum, J. M. Lee, G. R. Riley, W. Jang, W. S. Rubinstein, D. M. Church, D. R. Maglott, ClinVar: public archive of relationships among sequence variation and human phenotype. Nucleic Acids Res 42, D980–D985 (2014).

22. I. Adzhubei, D. M. Jordan, S. R. Sunyaev, Predicting functional effect of human missense mutations using PolyPhen-2. Curr Protoc Hum Genet Chapter 7, Unit7.20–Unit7.20 (2013).

23. N.-L. Sim, P. Kumar, J. Hu, S. Henikoff, G. Schneider, P. C. Ng, SIFT web server: predicting effects of amino acid substitutions on proteins. Nucleic Acids Res 40, W452–W457 (2012).

24. S. Bienert, A. Waterhouse, T. A. P. de Beer, G. Tauriello, G. Studer, L. Bordoli, T. Schwede, The SWISS-MODEL Repository—new features and functionality. Nucleic Acids Res 45, D313–D319 (2017).

25. I. Haq, G. E. Lowrey, N. Kalsheker, S. R. Johnson, Matrix metalloproteinase-12 (MMP-12) SNP affects MMP activity, lung macrophage infiltration and protects against emphysema in COPD. Thorax 66, 970–6 (2011).

26. N. Guyot, J. Wartelle, L. Malleret, A. A. Todorov, G. Devouassoux, Y. Pacheco, D. E. Jenne, A. Belaaouaj, Unopposed cathepsin G, neutrophil elastase, and proteinase 3 cause severe lung damage and emphysema. The American journal of pathology 184, 2197–210 (2014).

27. J. M. Shipley, R. L. Wesselschmidt, D. K. Kobayashi, T. J. Ley, S. D. Shapiro, Metalloelastase is required for macrophage-mediated proteolysis and matrix invasion in mice. Proceedings of the National Academy of Sciences 93, 3942–3946 (1996).

28. B. Suki, E. Bartolak-Suki, P. R. M. Rocco, Elastase-Induced Lung Emphysema Models in Mice. Methods Mol Biol. 1639 (2017), doi:10.1007/978-1-4939-7163-3_7.

29. J. M. D’Armiento, M. P. Goldklang, A. A. Hardigan, P. Geraghty, M. D. Roth, J. E. Connett, R. A. Wise, F. C. Sciurba, S. M. Scharf, J. Thankachen, M. Islam, A. J. Ghio, R. F. Foronjy, Increased Matrix Metalloproteinase (MMPs) Levels Do Not Predict Disease Severity or Progression in Emphysema. PLOS ONE 8, e56352 (2013).

30. I. K. Demedts, A. Morel-Montero, S. Lebecque, Y. Pacheco, D. Cataldo, G. F. Joos, R. A. Pauwels, G. G. Brusselle, Elevated MMP-12 protein levels in induced sputum from patients with COPD. Thorax 61, 196–201 (2006).

31. M. Ueno, T. Maeno, S. Nishimura, F. Ogata, H. Masubuchi, K. Hara, K. Yamaguchi, F. Aoki, T. Suga, R. Nagai, M. Kurabayashi, Alendronate inhalation ameliorates elastase-induced pulmonary emphysema in mice by induction of apoptosis of alveolar macrophages. Nat Commun 6, 6332 (2015).

32. A. Churg, S. Zhou, J. L. Wright, Matrix metalloproteinases in COPD. European Respiratory Journal 39, 197–209 (2012).

33. N. Ambalavanan, T. Nicola, P. Li, A. Bulger, J. Murphy-Ullrich, S. Oparil, Y. F. Chen, Role of matrix metalloproteinase-2 in newborn mouse lungs under hypoxic conditions. Pediatr Res 63, 26–32 (2008).

34. A. Chetty, G.-J. Cao, M. Severgnini, A. Simon, R. Warburton, H. C. Nielsen, Role of matrix metalloprotease-9 in hyperoxic injury in developing lung. Am J Physiol Lung Cell Mol Physiol 295, L584–L592 (2008).

35. A. Hilgendorff, K. Parai, R. Ertsey, G. Juliana Rey-Parra, B. Thebaud, R. Tamosiuniene, N. Jain, E. F. Navarro, B. C. Starcher, M. R. Nicolls, M. Rabinovitch, R. D. Bland, Neonatal mice genetically modified to express the elastase inhibitor elafin are protected against the adverse effects of mechanical ventilation on lung growth. American journal of physiology. Lung cellular and molecular physiology 303, L215–27 (2012).

36. C.-A. Brandsma, M. de Vries, R. Costa, R. R. Woldhuis, M. Königshoff, W. Timens, Lung ageing and COPD: is there a role for ageing in abnormal tissue repair? European Respiratory Review 26 (2017), doi:10.1183/16000617.0073-2017.

37. S. Sato, E. Bartolák-Suki, H. Parameswaran, H. Hamakawa, B. Suki, Scale dependence of structure-function relationship in the emphysematous mouse lung. Front Physiol 6, 146 (2015).

38. S. Liu, J. Cimprich, B. M. Varisco, Mouse pneumonectomy model of compensatory lung growth. Journal of visualized experiments : JoVE 1, 1–10 (2014).

39. M. S. Dunnill, Quantative Methods in the Study of Pulmonary Pathology. Thorax 17, 320–328 (1962).

40. S. M. Young, S. Liu, R. Joshi, M. R. Batie, M. Kofron, J. Guo, J. C. Woods, B. M. Varisco, Localization and stretch-dependence of lung elastase activity in development and compensatory growth. Journal of applied physiology (Bethesda, Md. : 1985) 118, 921–31 (2015).

41. M. Nagendran, D. P. Riordan, P. B. Harbury, T. J. Desai, Automated cell type classification in intact tissues by single-cell molecular profiling. eLife in revision (2017).

42. Y. Hao, S. Hao, E. Andersen-Nissen, W. M. Mauck, S. Zheng, A. Butler, M. J. Lee, A. J. Wilk, C. Darby, M. Zager, P. Hoffman, M. Stoeckius, E. Papalexi, E. P. Mimitou, J. Jain, A. Srivastava, T. Stuart, L. M. Fleming, B. Yeung, A. J. Rogers, J. M. McElrath, C. A. Blish, R. Gottardo, P. Smibert, R. Satija, Integrated analysis of multimodal single-cell data. Cell 184, 3573–3587.e29 (2021).

43. E. Y. Chen, C. M. Tan, Y. Kou, Q. Duan, Z. Wang, G. V. Meirelles, N. R. Clark, A. Ma’ayan, Enrichr: interactive and collaborative HTML5 gene list enrichment analysis tool. BMC Bioinformatics 14, 128 (2013).

44. R Core Team, R: A Language and Environment for Statistical Computing (R Foundation for Statistical Computing, Vienna, Austria, 2019; https://www.R-project.org/).

45. 45. A. Kassambara, ggpubr: Publication Ready Plots - Articles - STHDA (2020) (available at http://www.sthda.com/english/articles/24-ggpubr-publication-ready-plots/).

46. B. Auguie, gridExtra: Miscellaneous Functions for “Grid” Graphics (2017; https://CRAN.R-project.org/package=gridExtra).

47. C. O. Wilke, cowplot: Streamlined Plot Theme and Plot Annotations for “ggplot2” (2019; https://CRAN.R-project.org/package=cowplot).

48. G. Yu, ggplotify: Convert Plot to “grob” or “ggplot” Object (2019; https://CRAN.R-project.org/package=ggplotify).

49. T. Wei, V. Simko, R package “corrplot”: Visualization of a Correlation Matrix (2017; https://github.com/taiyun/corrplot).

50. A. Kassambara, rstatix: Pipe-Friendly Framework for Basic Statistical Tests (2020; https://CRAN.R-project.org/package=rstatix).

